# Environmental exposures are important risk factors for advanced liver fibrosis in African American adults: NHANES 1999-2018

**DOI:** 10.1101/2022.11.29.22282889

**Authors:** Ning Ma, Rowena Yip, Sara Lewis, Amreen Dinani, Christina Wyatt, Michael Crane, Artit Jirapatnakul, Li Li, Costica Aloman, Meena B. Bansal, Douglas Dieterich, Brooke Wyatt, David Yankelevitz, Claudia Henschke, Andrea D. Branch

## Abstract

**Background and aims:** The prevalence and etiology of liver fibrosis vary over time and impact racial/ethnic groups unevenly. This study measured time-trends and identified factors associated with advanced liver fibrosis in the U.S.

**Methods:** Standardized methods were used to analyze data on 47,422 participants (≥ 20 years) in the National Health and Nutrition Examination Survey (1999-2018). Advanced liver fibrosis was defined as Fibrosis-4 ≥2.67 and/or Forns Index ≥6.9 and elevated ALT.

**Results:** The estimated number of people with advanced liver fibrosis increased from 1.3 million (95% CI, 0.8-1.9) to 3.5 million (95% CI, 2.8-4.2), a nearly 3-fold increase. Prevalence was higher in non-Hispanic Black and Mexican American persons than in non-Hispanic White persons. In multivariable logistic regression analysis, cadmium was an independent risk factor in all racial/ethnic groups. Smoking and current excessive alcohol use were risk factors in most. Importantly, non-Hispanic Black persons had a distinctive set of risk factors compared to non-Hispanic White persons that included poverty (OR = 2.09; 95%CI, 1.44-3.03), and susceptibility to lead exposure (OR = 3.25; 95%CI, 1.95-5.43), but did not include diabetes (OR = 0.88; 95% CI, 0.61-1.27, P =0.52). Non-Hispanic Black persons were more likely to have high exposure to lead, cadmium, polychlorinated biphenyls, and poverty than Non-Hispanic White persons.

**Conclusions:** The number of people with advanced liver fibrosis has increased, creating a need to expand the liver care workforce. The risk factors for advanced fibrosis varied by racial/ethnicity. These variations provide useful information for the design of screening programs. Poverty and toxic exposures were associated with the high prevalence of advanced liver fibrosis in non-Hispanic Black persons and need to be addressed.

**Lay summary:** Because liver disease often produces few warning signs, simple and inexpensive screening tests that can be performed by non-specialists are needed to allow timely detection and linkage to care. This study shows that non-Hispanic Black persons have a distinctive set of risk factors that need to be taken into account when designing liver disease screening tests. Exposure to exogenous toxins may be especially important risk factors for advanced liver fibrosis in non-Hispanic Black persons.

## Introduction

Liver disease causes an estimated two million deaths globally each year^1^ but is under-diagnosed. In a recent U.S. study, nearly 50% of primary care patients who experienced a serious liver-related event did not have a prior diagnosis of liver disease.^2^ Similarly, in the United Kingdom, “around three-quarters of patients who will die from cirrhosis are currently unaware that they have liver disease”.^3^ Increased diagnosis will require screening for liver fibrosis in primary care settings.

The Fibrosis-4 (FIB-4) index is a validated marker of liver fibrosis (LF).^4, 5^ An American Gastroenterology Association task force reported that it provides a ‘useful, inexpensive, first-line assessment of liver fibrosis for use in primary care.”^5^ The Forns index is a second well-validated non-invasive fibrosis test.^6^ In the National Health and Nutrition Examination Survey (NHANES) population, FIB-4 ≥ 2.67 and Forns ≥ 6.9 have hazard ratios (HR)s for liver-related death of 42 and 117, respectively.^4^ When combined with alanine aminotransferase (ALT) elevation, both FIB-4 and Forns indices had areas under the receiver operating curve (AUROC) of 0.83 for predicting serious liver-related events over ten years in the community setting.^6^ Several additional screening tests have been proposed and it is likely that one or more will be broadly implemented soon.^6-8^

The American Association for the Study of Liver Diseases is currently developing guidelines for non-invasive LF screening.^9^ According to guidelines of the European Society for the Study of Liver (EASL),^10^ screening should be limited to individuals with known risk factors, such as type 2 diabetes. This restriction raises questions about which risk factors can be relied on to yield equitable and inclusive screening protocols.

Comprehensive information about risk factors in the multiethnic population of the United States (US) is lacking, but essential, particularly if screening is going to be limited to patients with specific risk factors. Given the high accuracy of FIB-4 and Forns indices when combined with elevated ALT to identify individuals at high risk for liver-related events,^6^ the objectives of this study are to use the nationally-representative NHANES data: a) to determine time-trends, b) to compare risk factors among racial/ethnic groups, and c) to determine the percentage of people with advanced LF who might be missed by etiology-based screening. The results show that the prevalence of advanced fibrosis nearly doubled over the past twenty years and was higher in Non-Hispanic Black (NHB) than in Non-Hispanic White (NHW) persons. NHB persons had a distinctive set of risk factors that included lead (Pb) and poverty but did not include diabetes or hypertension, which were risk factors in NHW persons. These differences need to be considered in risk factor-based screening guidelines. High cadmium exposure was a risk factor in all racial/ethnic groups, highlighting the potential role of environmental toxins in liver fibrosis.

## Materials and Methods

### Study population and data sources

NHANES uses standardized procedures to collect data under a protocol approved by the National Center for Health Statistics Research Ethnic Review Board. Analysis of de-identified NHANES data is exempt from IRB review.^11^ Ten cycles of NHANES (1999-2018) were used in the main analyses. Sub-studies used liver ultrasound data from NHANES III (1988-1994), and measurements of organic chemicals [polychlorinated biphenyls (PCB)] from NHANES 2003-2004. The public-use linked mortality file was obtained through 2019.^12^

### Indicators of fibrosis

FIB-4 and Forns indices were calculated as before.^4^ Advanced LF was indicated by FIB-4 ≥2.67 and/or Forns ≥6.9 and ALT above upper limit of normal (ULN) (≥40 IU/L for men, ≥31 IU/L for women).

### Demographic variables

Analysis used self-reported sex (male/female) and race/ethnicity [NHW, NHB, Mexican American (MA) and other (O) race (non-MA Hispanics and others)]. The main analysis was performed on people aged 20-85 years. Sensitivity analyses were conducted on people aged 35-64 because FIB-4 may under-estimate fibrosis in individuals younger than 35^13^ and because changes in health insurance may alter association with poverty after age 64.

### Definition of risk factors

Kidney insufficiency (KI) was a urinary albumin to creatinine ratio (UACR) ≥ 30mg/g and/or an estimated glomerular filtration rate (eGFR) < 60ml/min/1.73m^2^ calculated using the Chronic Kidney Disease Epidemiology Collaboration 2021 creatinine-based formula (race agnostic).^14^ Diabetes was self-reported, and/or hemoglobin A1c (HbA1c) ≥ 6.5%, and/or fasting plasma glucose (FPG) ≥ 126mg/dL.^15^ Hypertension was systolic blood pressure (SBP) ≥ 130mmHg, and/or diastolic blood pressure (DBP) ≥ 80mmHg, and/or use of anti-hypertensive medication.^16^ Body mass index (BMI) was categorized as normal weight (< 25Kg/m^2^), overweight (25 ≤ BMI < 30Kg/m^2^), and obese (≥ 30Kg/m^2^). Waist circumference (WC) was categorized as normal (< 94cm for men, < 80cm for women), moderate (94 ≤ WC < 102cm for men, 80 ≤ WC < 88cm for women) and high (≥ 102cm for men, ≥ 88cm for women).^17^ Metabolic syndrome was defined as ≥3 of the following: WC ≥ 102cm for men and ≥ 88cm for women; triglyceride ≥ 150mg/dL; HDL cholesterol < 40mg/dL for men and < 50mg/dL for women; SBP ≥ 130mmHg or DBP ≥ 85mmHg or taking hypertension medications; FPG ≥ 100mg/dL.^18^ Past/current smokers answered “Yes” to the question “Have you smoked at least 100 cigarettes in your lifetime?”, never smokers answered “No”.^19^ The responses to questions about alcohol consumption were used to create four mutually exclusive groups, lifetime abstainers (<12 drinks in lifetime), former drinkers (≥12 drinks in their lifetime but none in the past year), non-excessive current drinkers (on average, ≤14 drinks/week for men and ≤7 drinks/week for women, and never five or more in a single day during the past year for either), and excessive current drinkers (on average, >14 drinks/week for men and >7 drinks/week for women, or >5 drinks in a single day at least once during the past year for either).^20^ Blood levels of lead and cadmium were analyzed as continuous and binary variables [quartiles (Q)1-3 versus Q4]. Lipid-adjusted plasma levels of PCBs were classified by quartiles (Q1-3 versus Q4) (see Fig. S6 for details).^21^ Poverty was defined as a family poverty-income-ratio below 1.0.^22^

### Definitions of disease etiologies

Disease etiology was examined in participants who had data for calculating the US Fatty Liver Index (USFLI), which was previously validated for the US population.^23^ Viral hepatitis (VH) was past/current infection with hepatitis B virus (HBV), positive core antibody or surface antigen; or hepatitis C virus (HCV), RNA or antibody; Alcohol-associated liver disease (ALD) was meeting previous criteria^24^ and/or categorized as current excessive drinker^20^ in this study; Non-alcoholic fatty liver disease (NAFLD) was USFLI ≥30;^23^ and No Exposure Identified (NEI) was not meeting criteria for VH, ALD or NAFLD. In sensitivity analyses, NAFLD was defined by abdominal ultrasound (mild/moderate/severe fatty liver) from NHANES III.

### Statistical Analysis

All analysis were conducted according to NHANES guideline,^11^ using established methods to combine cycles. Data were adjusted for the complex NHANES design with strata, primary sampling units, and probability weights incorporated into statistical models using the survey estimation commands in SAS OnDemand for Academics (SAS Institute Inc., Cary, NC, USA). These procedures generate estimates for the housed, civilian, non-institutionalized population in the US. Age standardization estimates were calculated using the direct method, standardized to the 2000 US census population with four age categories for the 20-85 year age group and three age categories for the 35-64 year age group. Differences between groups were tested by univariate t statistics.^25^ To estimate the number of adults with advanced LF, prevalence was calculated and then multiplied by the estimated adult US population obtained from the Current Population Surveys or American Community Survey of each survey cycle.^26^ Annual percent changes (APC) were calculated using the Joinpoint Regression Program (Version 4.9.0.0, National Cancer Institute).^27^ Univariable and multivariable survey logistic regression with appropriate sample weights were used to examine the association between advanced LF and the independent variables. Survey-weighted adjusted multivariable cox proportional models were used to investigate the association between advanced-LF and all-cause mortality. Missing values that ranged from 0.1% to 9.0% and were addressed using multivariable imputation by chained equations.^28^ Combined estimates using ten imputed datasets were calculated. Statistical significance was a two-sided P value <0.05.

## Results

### Three-fold increase in cases of advanced LF over 20 years

The selection of the study group is presented in Fig. 1 and the dynamic time-trends of advanced LF are presented in Fig. 2. The estimated number of people with advanced LF increased from 1.3 million (95% CI, 0.8-1.9) to 3.5 million (95% CI, 2.8-4.2), a nearly 3-fold increase over 20 years (Fig. 2A). The age-standardized weighted prevalence approximately doubled (Fig. 2B). The annual percent change (APC) was 8.7% (95% CI, 6.7-10.9) (Table S1). The prevalence of advanced LF was about 1.6-fold higher in NHB than in NHW in the total group and in men and women when analyzed separately (Fig. 2C, D). The HR for all-cause mortality among people with advanced LF was 2.42 (95% CI, 1.96-2.97) in the 20-85 age group and 3.86 (95% CI, 2.78-5.37) in the 35-64 age group (Fig. 1B, C).

**Fig. 1.**
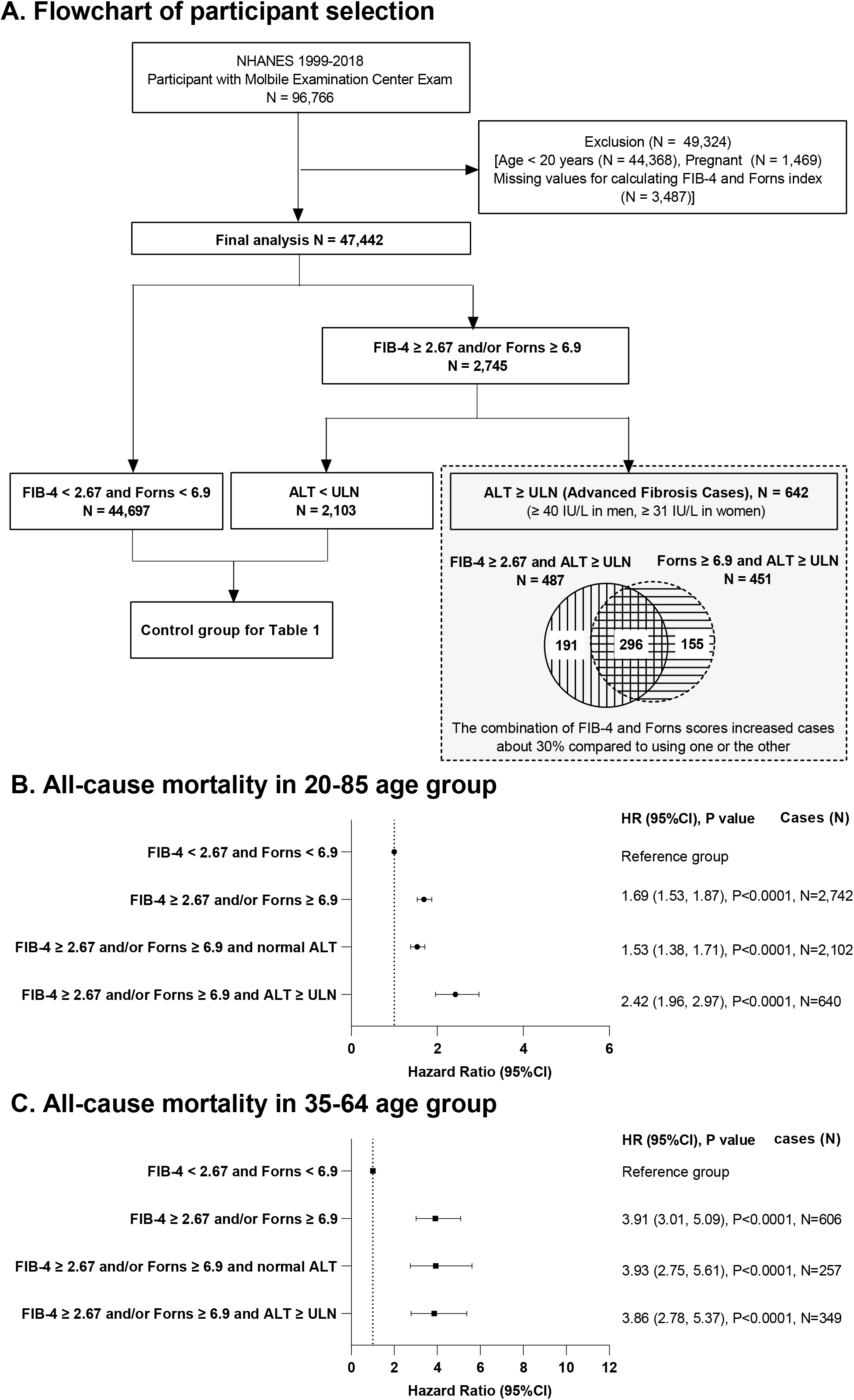
Flowchart of participant selection and association between advanced fibrosis and all-cause mortality. (A) Flowchart of participant selection; all-cause mortality (B) in the 20-85 age group and (C) in the 35-64 age group. Association between advanced fibrosis and all-cause mortality was analyzed using survey-weighted multivariable cox proportional models (adjusted for age, sex, race/ethnicity, body mass index, alcohol and smoking status, and poverty). ALT, alanine aminotransferase; FIB-4, fibrosis-4; NHANES, National Health and Nutrition Examination Survey; ULN, upper limit of normal.

**Fig. 2.**
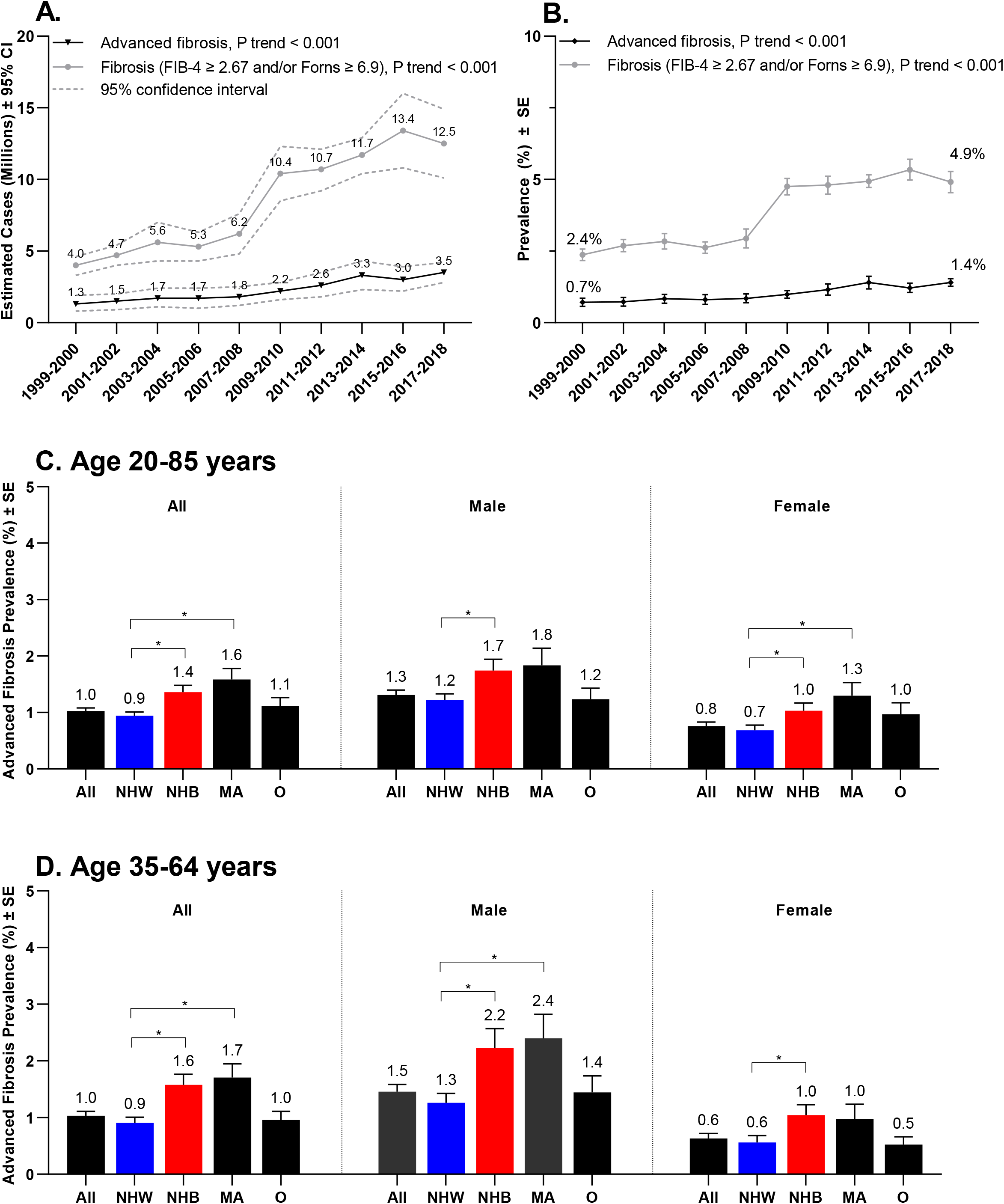
Time-trends and prevalence of advanced liver fibrosis. (A) The change in the number (millions) of U.S. adults with advanced liver fibrosis (black) and fibrosis defined by FIB-4 ≥2.67 and/or Forns ≥6.9 (grey) overtime with 95% confidence intervals (CI) (dashed); (B) age-standardized weighted prevalence of advanced fibrosis (black) and fibrosis (grey) over time. Age-standardized weighted prevalence of advanced fibrosis (C) in people 20-85 years old and (D) in people 35-64 years old (total, males, and females), stratified by race/ethnicity. Differences between groups were tested by univariate t statistic, *P<0.05. Abbreviations: Non-Hispanic White, NHW (blue); non-Hispanic Black, NHB (red); Mexican American, MA; other race, O; standard error, SE.

### Associated conditions

The age-standardized weighted prevalence of advanced LF was compared between people with and without health conditions that might be considered as eligibility criteria in risk factor-based screening. Several conditions differed by race/ethnicity. The prevalence of advanced LF was about 2-fold higher in those with diabetes in the total population and in NHW, MA and O, but not in NHB (total, males, and females) (Fig. 3A and Fig. S1,2); similar results were obtained when alternative definitions of diabetes were used, underscoring the robustness of the finding (Fig. S3). Only 35% (95% CI, 29.9-40.1) of participants with advanced LF had diabetes and thus 65% of cases would be missed if screening were limited to people with diabetes. Associations between obesity and advanced LF also differed by race/ethnicity. The prevalence of fibrosis was significantly higher in MA and O with obesity than in those with normal BMI (Fig. 3C). Strikingly, however, among NHB, the prevalence was about 2-fold higher in those with *<u>normal</u>* BMI than in those with obesity, with similar results were obtained for waist circumference (Fig. S4). Poverty was associated with advanced LF in NHB, but not in any other racial/ethnic group. Of the six conditions that reflect exposure to exogenous toxins, four (smoking, current excessive drinking, cadmium exposure, and lead exposure) were associated with advanced LF in the total population, in NHW, and in NHB (Fig. 3F, I, K, L). Former drinkers had a higher prevalence of advanced LF in NHB (Fig. 3G). In sensitivity analyses, similar results were obtained when a less restrictive definition of LF (without the requirement for ALT elevation) was used (Fig. S5).

**Fig. 3.**
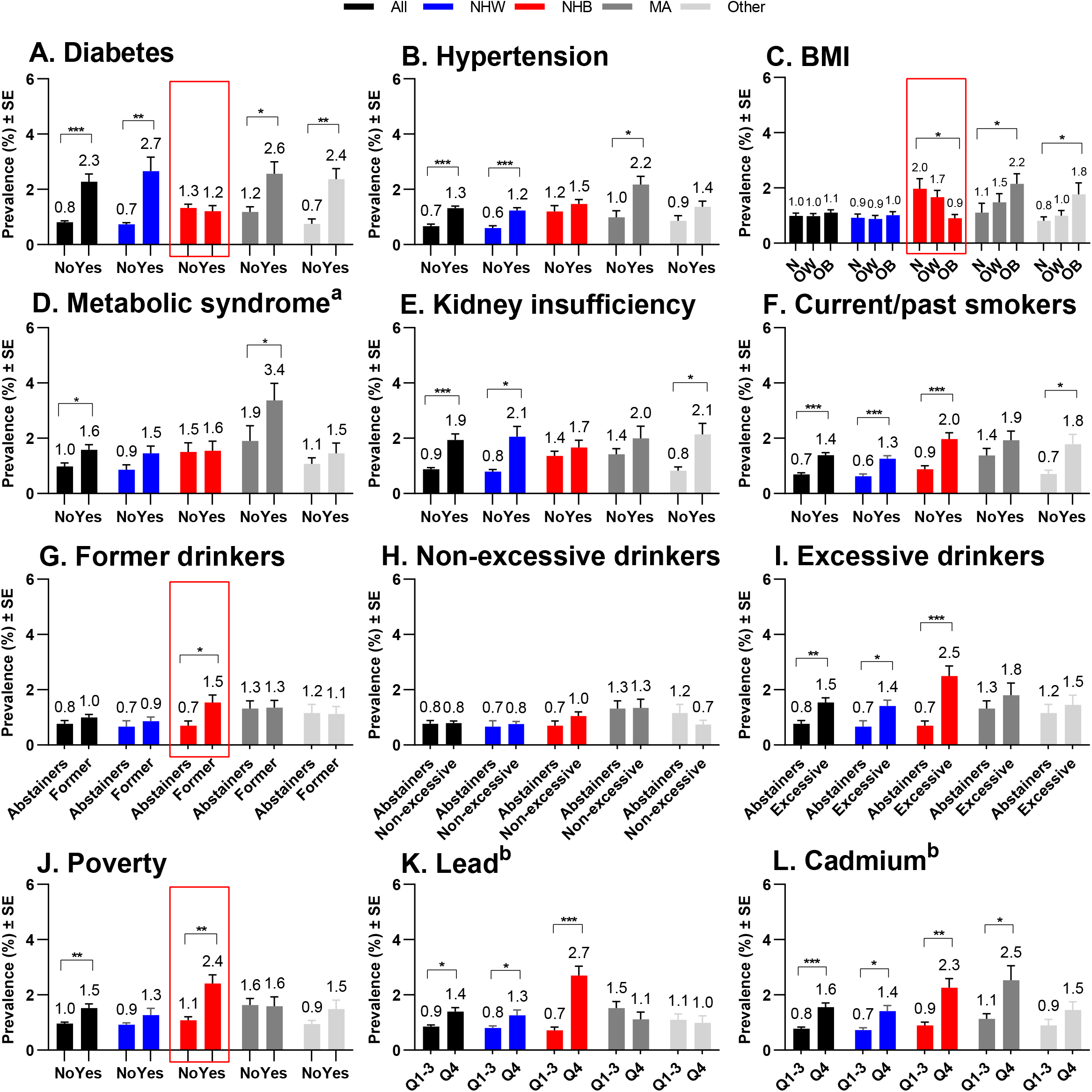
Advanced fibrosis in participants aged 20-85 years, stratified by health conditions and race/ethnicity. The age-standardized weighted prevalence of advanced fibrosis among participants with and without various health conditions were determined for the total cohort (black), non-Hispanic White (NHW, blue), non-Hispanic Black (NHB, red), Mexican American (MA, grey) and other (light grey) racial/ethnic groups. (A) Diabetes, (B) hypertension, (C) body mass index (BMI) categories, normal (N), overweight (OW) and obese (OB), (D) metabolic syndrome, (E) kidney insufficiency, (F) current/past smokers, (G) former drinkers vs. lifetime abstainers, (H) current non-excessive drinkers vs. lifetime abstainers, (I) current excessive drinkers vs. lifetime abstainers, (J) poverty, (K) blood levels of lead and (L) cadmium (Q1-3 vs. Q4). Differences between groups were tested by univariate t statistic. *P < 0.05, **P < 0.001, ***P < 0.0001. ^a^ Components of metabolic syndrome were only available for participants with fasting blood tests in NHANES 2007-2018, N=13,886. ^b^ Cadmium and lead analysis were based on complete datasets without imputation, N=42,255. Abbreviation: quartile, Q; standard error = SE.

### Prevalence of risk factors

Compared to NHW, NHB had a lower prevalence of smoking and excessive drinking; however, they had a higher prevalence of many other conditions, including diabetes. Thus, the disconnection between diabetes and advanced LF in NHB does not result from a low prevalence of diabetes. NHB also had a higher prevalence of KI, hypertension, obesity, poverty, and exposure to environmental pollutants, as indicated by higher blood levels of lead, cadmium, and PCBs (Fig. 4). Both heavy metals and organic chemicals are associated with liver disease.^21, 29, 30^

**Fig. 4.**
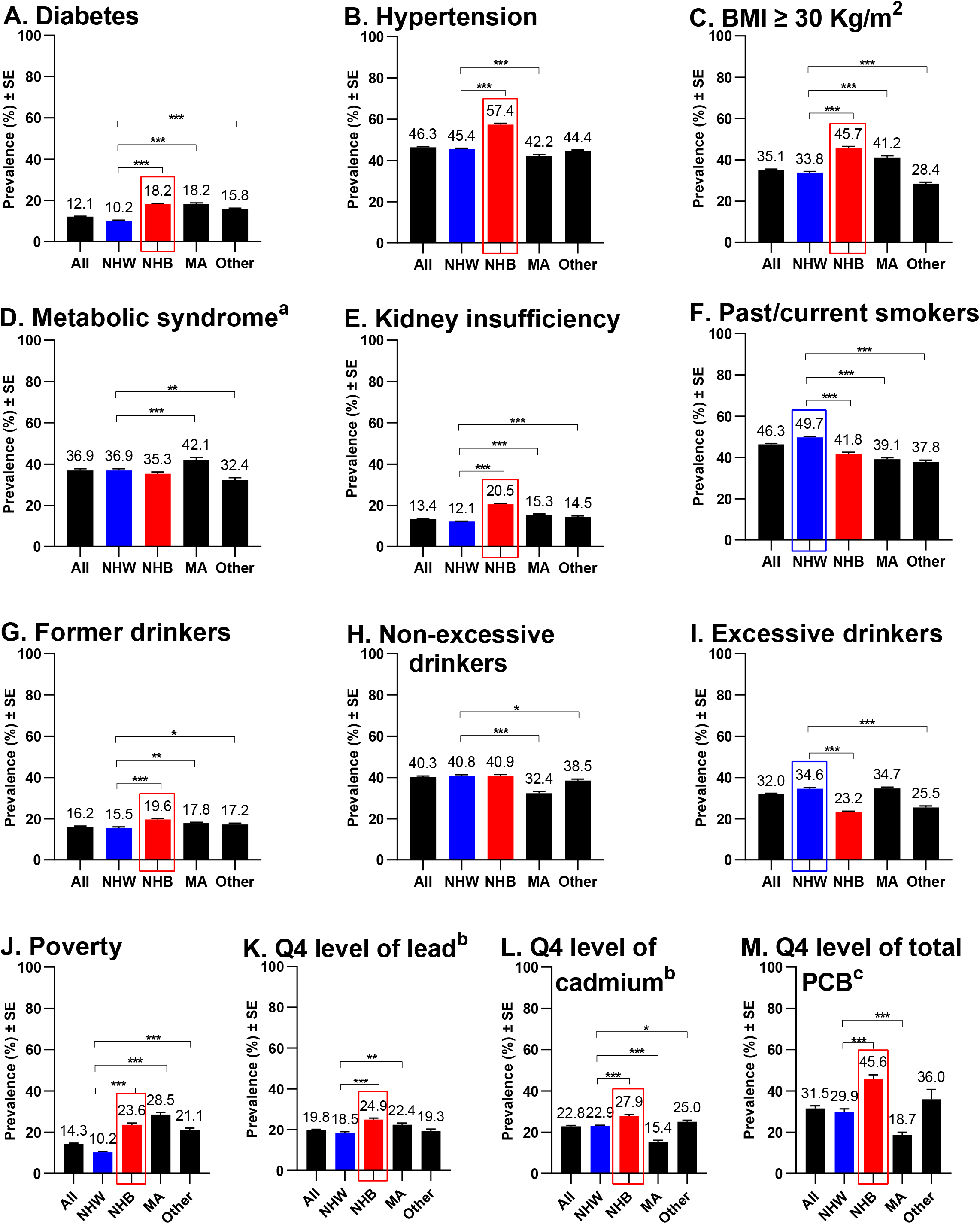
Prevalence of heath conditions in participants aged 20-85 years, stratified by race/ethnicity. Shown are the age-standardized weighted prevalence of (A) diabetes, (B) hypertension, (C) obesity (BMI ≥ 30 Kg/m2), (D) metabolic syndrome, (E) kidney insufficiency, (F) past/current smokers, (G) former drinker, (H) current non-excessive drinker, (I) current excessive drinker, (J) poverty, quartile 4 (Q4) blood level of (K) lead, (L) cadmium, and (M) total polychlorinated bi-phenyls (PCB) among non-Hispanic White (NHW, blue), non-Hispanic Black (NHB, red), Mexican American (MA), and Other race groups. ^a^ Components of metabolic syndrome were only available for participants with fasting blood test in NHANES 2007-2018, N=13,886. ^b^ Cadmium and lead analysis were based on complete datasets without imputation, N=42,255. ^c^ PCB data from NHANES 2003-2004 survey cycles included 1,242 participants. Differences between groups were tested by univariate t statistic, *P < 0.05, **P < 0.001, ***P < 0.0001. Abbreviation: body mass index, BMI; standard error, SE.

### Conditions independently associated with advanced LF

Multivariable logistic regression was used to identify factors independently associated with advanced LF. Variables included age, sex, KI, diabetes, hypertension, BMI, alcohol use, smoking, and poverty. Two age groups were analyzed (20-85 and 36-64 years). In the 20-85 age group, smoking and current excessive drinking were risk factors in both NHW and NHB (Table 1). Diabetes and hypertension were independently associated with advanced LF in NHW, but not in NHB. Conversely, poverty was a risk factor in NHB, but not in NHW. In a sensitivity analysis that excluded participants with viral hepatitis, generally similar odds ratios (OR)s obtained; among NHB, the OR for diabetes was 0.76 (95% CI, 0.39-1.48) and the OR for smoking 3.00 (95% CI, 1.56-5.78) (Table S2). When metabolic syndrome was included (rather than diabetes, hypertension, and obesity), it was associated with advanced LF in the total group (Table S3). Results for the 20-85 and the 35-64 age groups were generally similar; however, in the 35-64 age group, KI was a risk factor for advanced LF in NHW; and poverty was a risk factor in NHW, as well as in NHB (Table S4). KI was an independent risk factor in all racial/ethnic groups in a sensitivity analysis that used the less restrictive definition of LF (Table S5).

**Table 1.**
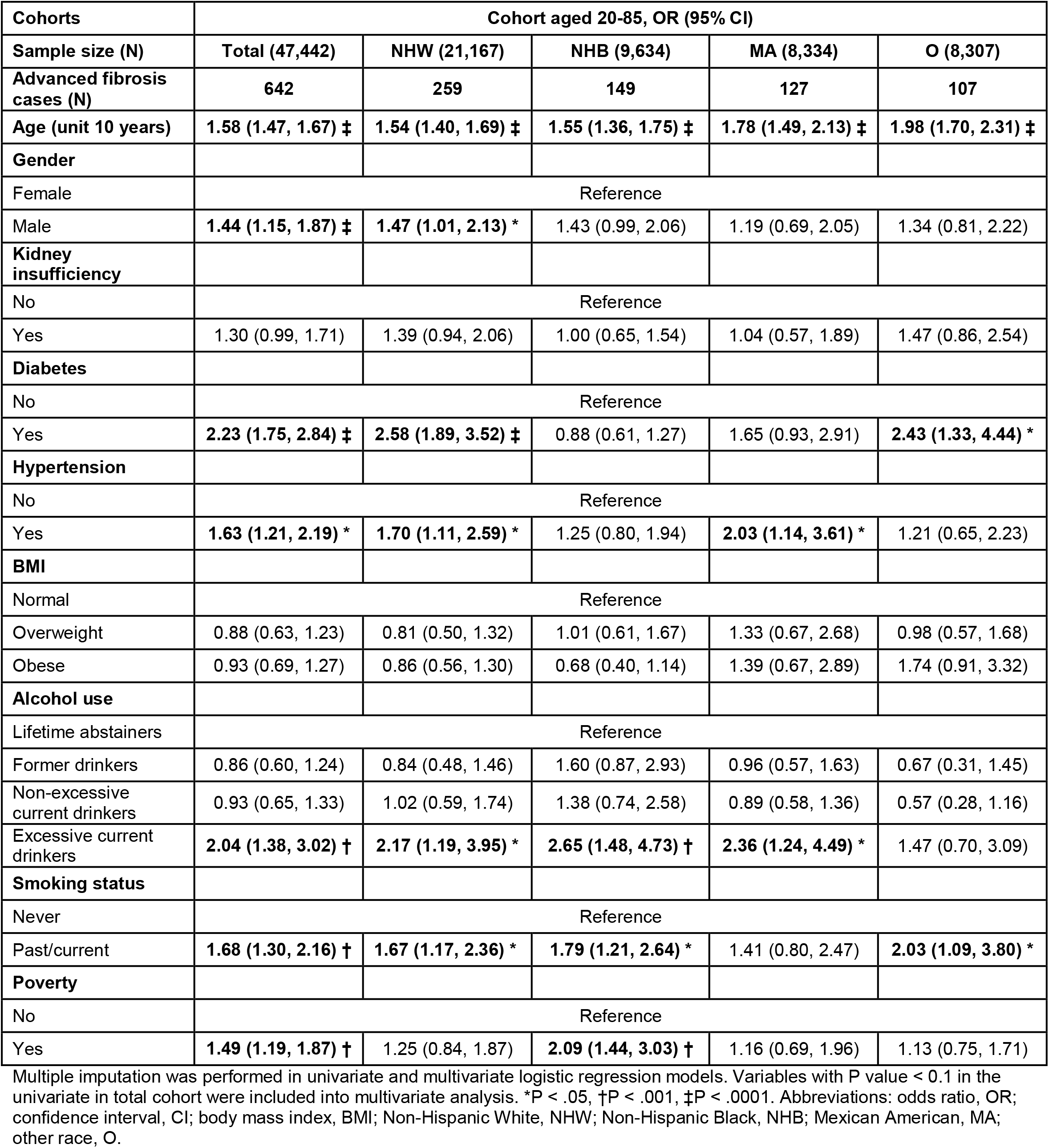
Odds ratios from multivariable logistic regression models with the outcome of advanced liver fibrosis.

### Environmental exposures and advanced LF

High blood levels of cadmium, as indicated by measurements in the 4^th^ quartile (Q4), were associated with advanced LF in the total population and in NHW, NHB, and MA in multivariable logistic regression analysis (Table 2). Blood levels of lead were strongly associated with advanced LF in NHB, but not in NHW: OR = 3.25 (95% CI, 1.95-5.43) vs. 1.24 (95% CI, 0.79-1.94, P=0.34) (Table 3). Associations between advanced LF and heavy metal exposures were dose-dependent for both cadmium and lead (Table S6,7).

**Table 2.** Odds ratios from multivariable logistic regression models with the outcome of advanced liver fibrosis and added blood cadmium level into models.

| Cohorts | In aged 20-85 years with cadmium into models, OR (95% CI) |  |  |  |  |
| --- | --- | --- | --- | --- | --- |
| Sample size (N) | Total (42,255) | NHW (19,176) | NHB (8,585) | MA (7,508) | O (6,986) |
| Advanced fibrosis cases (N) | 542 | 226 | 128 | 102 | 86 |
| Survey year | 1.07 (1.03, 1.11) ‡ | 1.08 (1.03, 1.13) † | 1.01 (0.95, 1.08) | 1.10 (0.99, 1.20) | 1.03 (0.94, 1.13) |
| Age (unit 10 years) | 1.56 (1.44, 1.68) ‡ | 1.51 (1.36, 1.69) ‡ | 1.52 (1.31, 1.76) ‡ | 1.79 (1.46, 2.21) ‡ | 2.09 (1.75, 2.50) ‡ |
| Gender |  |  |  |  |  |
| Female | Reference |  |  |  |  |
| Male | 1.61 (1.24, 2.08) † | 1.67 (1.16, 2.41) * | 1.61 (1.08, 2.40) * | 1.30 (0.71, 2.36) | 1.25 (0.65, 2.40) |
| Kidney insufficiency |  |  |  |  |  |
| No | Reference |  |  |  |  |
| Yes | 1.15 (0.85, 1.55) | 1.16 (0.75, 1.79) | 1.17 (0.74, 1.84) | 0.58 (0.36, 0.94) * | 1.54 (0.83, 2.87) |
| Diabetes |  |  |  |  |  |
| No | Reference |  |  |  |  |
| Yes | 2.38 (1.83, 3.11) ‡ | 2.85 (2.05, 3.96) ‡ | 0.78 (0.50, 1.19) | 1.97 (1.04, 3.73) * | 2.60 (1.26, 5.35) * |
| Hypertension |  |  |  |  |  |
| No | Reference |  |  |  |  |
| Yes | 1.86 (1.33, 2.58) † | 2.04 (1.27, 3.27) * | 1.32 (0.77, 2.24) | 2.37 (1.15, 4.86) * | 1.13 (0.54, 2.35) |
| BMI |  |  |  |  |  |
| Normal | Reference |  |  |  |  |
| Overweight | 0.89 (0.60, 1.33) | 0.85 (0.48, 1.49) | 0.91 (0.52, 1.57) | 1.15 (0.56, 2.34) | 0.94 (0.43, 2.08) |
| Obese | 0.92 (0.64, 1.32) | 0.81 (0.49, 1.33) | 0.80 (0.47, 1.39) | 1.02 (0.46, 2.30) | 2.00 (0.83, 4.85) |
| Alcohol use |  |  |  |  |  |
| Lifetime abstainers | Reference |  |  |  |  |
| Former drinkers | 0.92 (0.61, 1.38) | 0.85 (0.49, 1.47) | 1.91 (0.86, 4.25) | 0.84 (0.30, 2.35) | 0.87 (0.33, 2.29) |
| Non-excessive current drinkers | 1.00 (0.66, 1.52) | 1.06 (0.61, 1.85) | 1.41 (0.59, 3.34) | 0.81 (0.26, 2.50) | 0.74 (0.30, 1.82) |
| Excessive current drinkers | 2.02 (1.30, 3.13) * | 2.08 (1.12, 3.85) * | 2.65 (1.19, 5.90) * | 2.03 (0.63, 6.46) | 1.70 (0.64, 4.57) |
| Smoking status |  |  |  |  |  |
| Never | Reference |  |  |  |  |
| Past/current | 1.40 (1.03, 1.91) * | 1.42 (0.94, 2.14) | 1.23 (0.76, 2.00) | 1.37 (0.67, 2.80) | 1.71 (0.72, 4.06) |
| Poverty |  |  |  |  |  |
| No | Reference |  |  |  |  |
| Yes | 1.36 (1.02, 1.80) * | 1.00 (0.62, 1.62) | 1.95 (1.26, 3.02) * | 1.11 (0.57, 2.19) | 1.06 (0.56, 2.01) |
| Blood cadmium level |  |  |  |  |  |
| Q1-3 | Reference |  |  |  |  |
| Q4 | 1.81 (1.30, 2.53) † | 1.75 (1.08, 2.85) * | 2.01 (1.23, 3.30) * | 2.24 (1.21, 4.16) * | 1.75 (0.83, 3.65) |

Cadmium analysis based on complete dataset with information of blood lead and cadmium measurements with N=42,255. Multiple imputation was performed in univariate and multivariate logistic regression models. Variables with P value < 0.1 in the univariate in total cohort were included into multivariate analysis. \*P < 0.05, †P < 0.001, ‡P < 0.0001. Abbreviations: odds ratio, OR; confidence interval, CI; body mass index, BMI; Non-Hispanic White, NHW; Non-Hispanic Black, NHB; Mexican American, MA; other race, O.

**Table 3.** Odds ratios from multivariable logistic regression models with the outcome of advanced liver fibrosis and added blood lead level into models.

| Cohorts | In aged 20-85 years with blood lead (Pb) level into models, OR (95% CI) |  |  |  |  |
| --- | --- | --- | --- | --- | --- |
| Sample size (N) | Total (42,255) | NHW (19,176) | NHB (8,585) | MA (7,508) | O (6,986) |
| Advanced fibrosis cases (N) | 542 | 226 | 128 | 102 | 86 |
| Survey year | 1.08 (1.04, 1.12) ‡ | 1.09 (1.04, 1.14) ‡ | 1.07 (1.01, 1.14) * | 1.06 (0.97, 1.17) | 1.02 (0.92, 1.12) |
| Age (unit 10 years) | 1.52 (1.40, 1.65) ‡ | 1.47 (1.31, 1.66) ‡ | 1.35 (1.16, 1.59) ‡ | 1.83 (1.49, 2.26) ‡ | 2.12 (1.78, 2.53) ‡ |
| Gender |  |  |  |  |  |
| Female | Reference |  |  |  |  |
| Male | 1.46 (1.11, 1.92) * | 1.54 (1.04, 2.28) * | 1.12 (0.72, 1.74) | 1.24 (0.69, 2.22) | 1.19 (0.67, 2.14) |
| Kidney insufficiency |  |  |  |  |  |
| No | Reference |  |  |  |  |
| Yes | 1.19 (0.88, 1.59) | 1.20 (0.79, 1.84) | 1.01 (0.46, 2.24) | 0.63 (0.38, 1.05) | 1.58 (0.85, 2.93) |
| Diabetes |  |  |  |  |  |
| No | Reference |  |  |  |  |
| Yes | 2.37 (1.82, 3.10) ‡ | 2.84 (2.04, 3.95) ‡ | 0.82 (0.53, 1.26) | 1.85 (0.99, 3.44) | 2.56 (1.27, 5.14) † |
| Hypertension |  |  |  |  |  |
| No | Reference |  |  |  |  |
| Yes | 1.87 (1.34, 2.60) † | 2.04 (1.27, 3.29) * | 1.29 (0.76, 2.20) | 2.38 (1.16, 4.89) * | 1.12 (0.53, 2.35) |
| BMI |  |  |  |  |  |
| Normal | Reference |  |  |  |  |
| Overweight | 0.85 (0.58, 1.25) | 0.81 (0.47, 1.42) | 0.89 (0.51, 1.56) | 1.08 (0.51, 2.28) | 0.87 (0.41, 1.87) |
| Obese | 0.86 (0.61, 1.21) | 0.76 (0.47, 1.22) | 0.83 (0.48, 1.44) | 0.93 (0.42, 2.09) | 1.73 (0.70, 4.27) |
| Alcohol use |  |  |  |  |  |
| Lifetime abstainers | Reference |  |  |  |  |
| Former drinkers | 0.91 (0.61, 1.37) | 0.87 (0.50, 1.50) | 1.72 (0.77, 3.87) | 0.84 (0.30, 2.30) | 0.86 (0.34, 2.14) |
| Non-excessive current drinkers | 0.97 (0.64, 1.46) | 1.05 (0.61, 1.81) | 1.33 (0.56, 3.14) | 0.78 (0.25, 2.41) | 0.75 (0.31, 1.77) |
| Excessive current drinkers | 1.96 (1.26, 3.05) * | 2.05 (1.10, 3.81) * | 2.38 (1.08, 5.25) * | 2.07 (0.65, 6.55) | 1.71 (0.66, 4.43) |
| Smoking status |  |  |  |  |  |
| Never | Reference |  |  |  |  |
| Past/current | 1.68 (1.27, 2.22) † | 1.68 (1.15, 2.45) * | 1.44 (0.92, 2.25) | 1.75 (0.92, 3.33) | 1.96 (0.90, 4.24) |
| Poverty |  |  |  |  |  |
| No | Reference |  |  |  |  |
| Yes | 1.36 (1.02, 1.80) * | 1.07 (0.66, 1.73) | 1.86 (1.19, 2.90) * | 1.21 (0.64, 2.31) | 1.10 (0.59, 2.06) |
| Blood lead level |  |  |  |  |  |
| Q1-3 | Reference |  |  |  |  |
| Q4 | 1.32 (0.96, 1.80) | 1.24 (0.79, 1.94) | 3.25 (1.95, 5.43) ‡ | 0.72 (0.40, 1.28) | 0.84 (0.39, 1.83) |

Lead analysis based on complete dataset with information of blood lead and cadmium measurements with N=42,255. Multiple imputation was performed in univariate and multivariate logistic regression models. Variables with P value < 0.1 in the univariate in total cohort were included into multivariate analysis. \*P < 0.05, †P < 0.001, ‡P < 0.0001. Abbreviations: odds ratio, OR; confidence interval, CI; body mass index, BMI; Non-Hispanic White, NHW; Non-Hispanic Black, NHB; Mexican American, MA; other race, O.

NHANES 2003-2004 measured lipid-adjusted plasma levels of PCBs in 1,242 adults (Fig. S6). Over 95% of the participants with advanced LF had high PCB exposure (Fig. 5). The small sample size precluded an analysis by race/ethnicity.

**Fig. 5.**
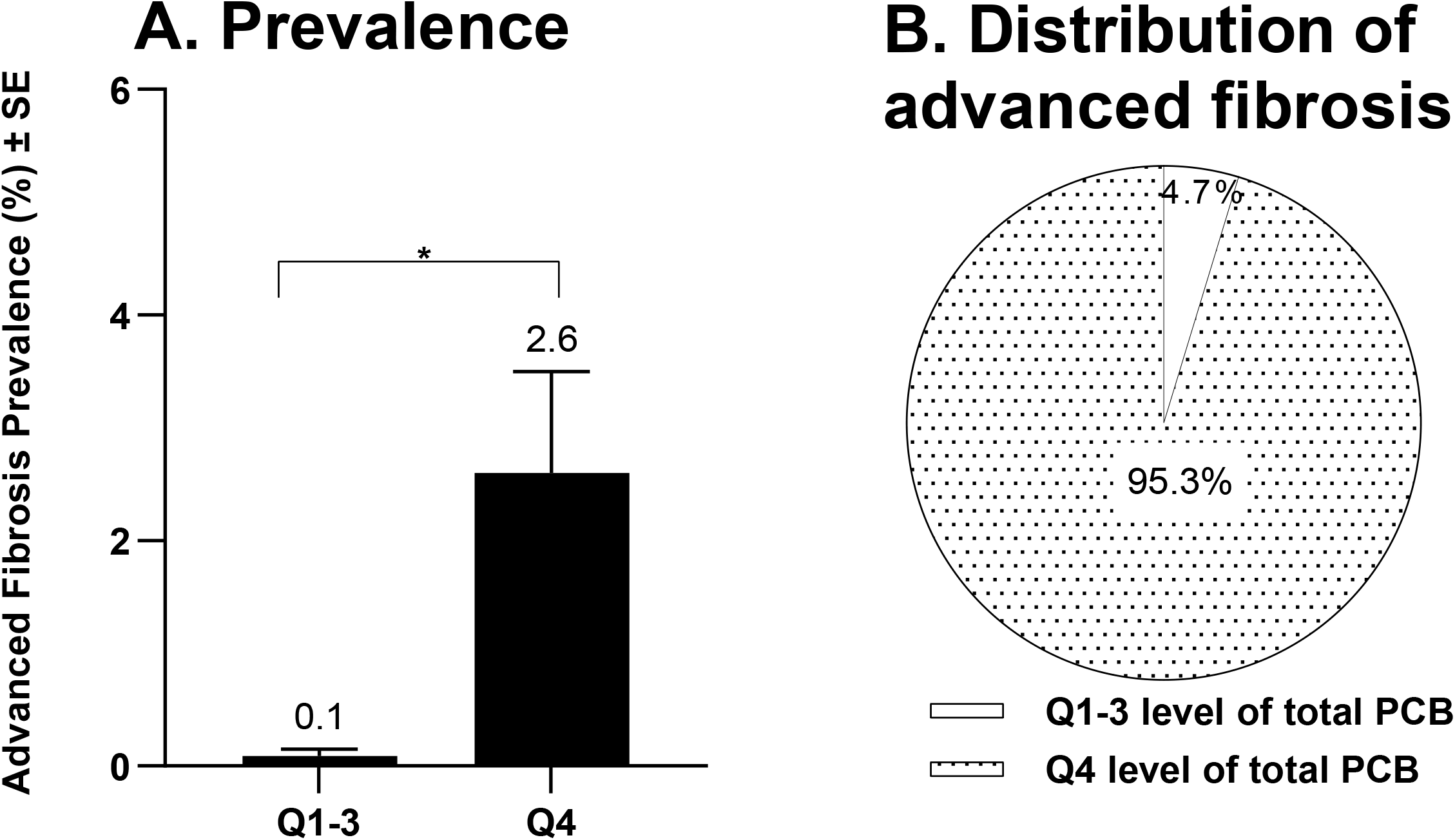
Association between advanced fibrosis and polychlorinated biphenyls (NHANES 2003-2004). (A) the age-standardized weighted prevalence of advanced liver fibrosis in people with low [quartile (Q)1-3] and high (Q4) lipid-adjusted plasma measurements of total PCBs in participants in NHANES 2003-2004. (B) distribution of advanced fibrosis cases in participants with Q1-3 (white) versus Q4 (dotted) levels of total PCBs. Differences between groups were tested by univariate t statistic, *P<0.05.

### Time-trends of health conditions from 1999-2000 to 2017-2018

To identify factors that might underlie the increase in advanced LF and to determine whether the 8.7% APC was typical of other diseases, we examined time-trends for 13 other conditions (Table S1). The age-standardized-weighted prevalence of high lead and cadmium exposure decreased, as did the percentage of former drinkers in the total population, in NHW, and in NHB. Smoking decreased in the total population, but the decrease was not significant among NHB. KI, hypertension, and current non-excessive drinking did not change significantly. Diabetes increased 1.6-fold in the total population (APC=4.9%) and in all groups except NHB. Obesity, as defined by either BMI or WC, increased; however, obesity was not associated with advanced LF in multivariable logistic regression analyses. LF without the requirement for ALT elevation increased 2.0-fold in the total population (APC=10.7%), suggesting that advanced LF may continue to rise in the future (Table S1). Current excessive drinking increased significantly in the total population (APC=2.3%) and increased 1.8-fold in NHB (APC=5.8%), underscoring the importance of alcohol in liver fibrosis.

### Fibrosis in adults with no exposure identified (NEI)

Screening is often used to detect patients with *specific* liver diseases. We investigated the percentage of people with advanced LF who might be missed if the population were screened for the three major liver diseases, VH, ALD, and NAFLD, rather than for fibrosis. One analysis used the USFLI to define NAFLD (n=20,388). Among the 285 with advanced LF, 36 (12.8%, 95% CI, 7.5-18.0) did not meet criteria for VH, ALD, or NAFLD. A second analysis used ultrasound to define NAFLD (n=12,811 NHANES III participants). Among the 84 with advanced LF, 10 (13.9%, 95% CI, 3.6-24.2) did not meet criteria for VH, ALD, or NAFLD (Fig. S7,8). In sensitivity analyses that used LF without the requirement for ALT elevation, almost 40% of the cases were in the no exposure identified (NEI) category (Fig. S9,10). This corresponds to 3.24 million U.S. adults averaged over the time-period analyzed. Multivariable logistic regression identified KI as a risk factor for LF in the NEI category (Table S8). These findings suggest that a significant percentage of advanced LF occurs in individuals who could be missed in etiology-based screening programs.

## Discussion

This study used nationally representative data to evaluate dynamic changes in the prevalence of advanced LF and to identify risk factors in the multi-ethnic U.S. population. The results provide valuable information for the design of liver disease screening tests. The study had three major findings.

First, during the past 20 years, the prevalence of advanced LF approximately doubled and increased more rapidly than 13 other conditions. The number of people with advanced LF increased nearly 3-fold, reaching about 3.5 million in the 2017-2018 NHANES cycle. Liver services will need to expand to care for these patients. Only diabetes increased more than 1.5-fold in the total population (APC, 4.9%), which makes the increase in advanced LF (APC, 8.7%) especially noteworthy. While diabetes rose among NHW, it did not increase significantly in NHB. Conversely, current excessive drinking increased 1.8-fold (APC, 5.8%) among NHB and may be an important driver. Excessive current drinking increased about 1.2-fold in the total population (APC, 2.3%), underscoring the need to reduce harmful drinking.

Second, advanced LF was strongly associated with heavy metal (lead and cadmium) exposure and over 95% of participants with advanced LF had high lipid-adjusted levels of PCBs. These findings add to published data^21, 29-33^ and should prompt a more extensive examination of toxic exposures in liver disease. Importantly, the World Health Organization classifies cadmium as a known human carcinogen.^34^ Additional factors independently associated with advanced LF were older age, male sex, diabetes, hypertension, excessive current drinking, past/current smoking, and poverty. Kidney insufficiency was independently associated with advanced LF in the 35-64 year age group, consistent with previous reports.^35, 36^ The association between LF and KI may reflect the shared roles of the liver and kidney in metabolism, detoxification, and excretion.

Third, NHB (both men and women) had a higher prevalence of advanced LF than their NHW counterparts and different risk factors. These results add to past evidence that NHB have a distinctive pattern of liver disease presentation and genomic factors.^32, 37-40^ A previous analysis of NHANES data also showed that NHB have a higher prevalence of cirrhosis,^41^ while, other studies reported a lower prevalence of biopsy-defined advanced LF among NHB.^42^ Because NHB are often under-represented in clinical trials,^43^ and may have incomplete medical records and less access to healthcare,^44^ nationally representative samples, as provided by NHANES, are especially important. Diabetes was independently associated with advanced LF in NHW^45^ and O, but not in NHB, as shown before,^45^ or in MA, which is consistent with published data, as past studies did not adjust for hypertension and kidney insufficiency.^46^ Among NHB, high blood levels of lead were strongly associated with advanced liver fibrosis (OR=3.25) and poverty was also a risk factor. Poverty is associated with workplace and environmental toxic exposures.^47^ High blood levels of PCBs were strongly associated with advanced LF and NHB had higher blood levels than NHW. Compared to NHW, NHB have a higher prevalence of a polymorphism in the gene encoding arylsulfatase A, a metabolic regulator^32^ associated with neurotoxicity,^33^ and they develop lung cancer at younger ages and with fewer pack-years of smoking,^31^ suggesting they may be especially vulnerable to toxic injury. In this study, NHB had a higher prevalence of KI, hypertension, obesity, poverty, and exposure to environmental pollutants (lead, cadmium, and PCB). These disparities could be associated with their reduced longevity.^48^

The study provided intriguing evidence that even if everyone in the U.S. were fully screened for VH, ALD, and NAFLD (don’t hold your breath), 12-40% of significant LF might be missed. These findings are consistent with data showing that about 20% of cirrhosis-related deaths occur in people without any of the major liver diseases,^49^ and with results showing that liver disease etiology was unspecified in 48% of cirrhosis- or hepatocellular carcinoma-related deaths in the U.S.^50^ These findings highlight the advantage of universal screening for advanced liver fibrosis. At a negligible cost, electronic health records could flag patients with FIB-4 ≥2.67 and/or Forns ≥6.9 and ALT ≥ULN, providing a realistic backstop to risk factor-based and etiology-based screening and offering a safety net for the high percentage of non-diabetics whose liver disease has not been diagnosed.

## Limitations

The main limitations are: a) the use of NHANES data, which are collected cross-sectionally at a single time point and are restricted to the housed non-institutionalized population; b) the use of the FIB-4/Forns scores and USFLI to define LF and NAFLD, rather than histopathology; c) the use of self-reported data to define race/ethnicity, alcohol use, and smoking habits. The study could not assess causality. To mitigate these limitations, we a) acknowledge them here, b) performed weighted and age-standardized analyses, which adjust for changes in the age and demographic structure of the population, and c) used ultrasound to define NAFLD in confirmatory studies.

## Conclusions

In the U.S., the prevalence of advanced LF doubled over the past 20 years and was higher in NHB (total group and men and women) than in NHW (total group and men and women). Liver care services will need to expand to meet the increased liver disease burden. Toxic exposures had especially strong associations with LF in NHB, suggesting that NHB may be particularly vulnerable. Poverty, smoking, excessive drinking, and exposure to environmental toxins are potentially modifiable LF risk factors. Universal screening with FIB-4 ≥2.67 and/or Forns ≥6.9 and ALT ≥ULN would be a realistic backstop to risk factor-based screening.

## Data Availability

All data produced are available online at https://wwwn.cdc.gov/nchs/nhanes/default.aspx

## Abbreviations

(ALD): Alcohol-associated liver disease
(ALT): Alanine aminotransferase
(AUROC): Area under the receiver operating curve
(BMI): Body mass index
(CI): Confidence interval
(KI): Fibrosis-4
(FIB-4): score; Kidney insufficiency
(HR): Hazard ratio
(LF): Liver fibrosis
(MA): Mexican American
(NHW): Non-Hispanic White
(NHB): Non-Hispanic Black
(NAFLD): Non-alcoholic fatty liver disease
(NHANES): National Health and Nutrition Evaluation Survey
(NEI): No exposure identified
(PCB): Polychlorinated biphenyls
(Q): Quartile
(VH): Viral hepatitis

## Acknowledgements

The authors thank Dr. Kerry Willis (National Kidney Foundation) for highlighting the likely importance of kidney insufficiency. This work was supported by National Institute for Occupational Safety and Health (NIOSH): U01 OH01163.

**Supplementary material is available on the preprint site**.

Author names in bold designate shared co-first authorship

