## Supplementary material for "Environmental exposures are important risk factors for advanced liver fibrosis in African American adults: NHANES 1999-2018"

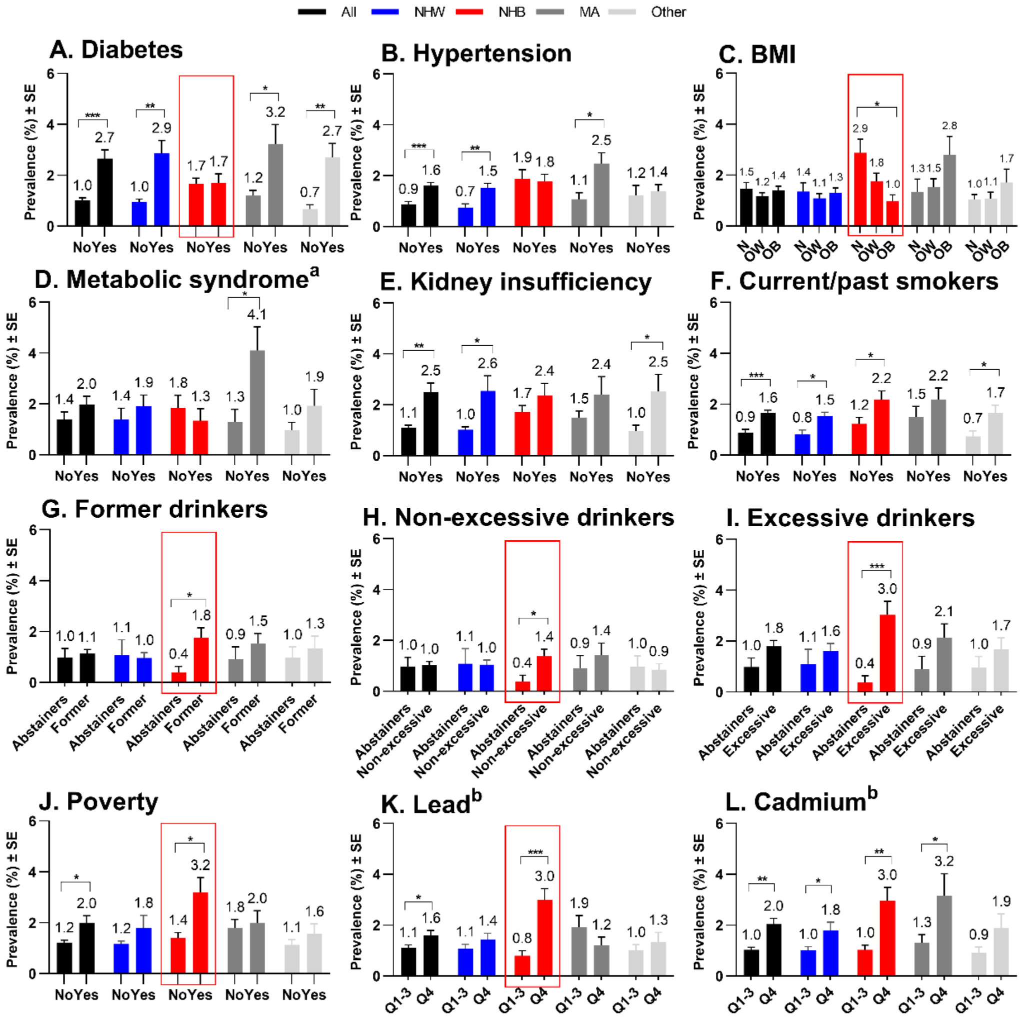

**Fig. S1. Age standardized weighted prevalence of advanced fibrosis in _**males**_ aged 20-85 years, stratified by health conditions and race/ethnicity.** The age-standardized weighted prevalence of advanced fibrosis among male participants with and without various health conditions were determined for the total cohort (black), non-Hispanic White (NHW, blue), non-Hispanic Black (NHB, red), Mexican American (MA, grey) and other (light grey) racial/ethnic groups. (A) Diabetes (Yes/No), (B) hypertension (Yes/No), (C) body mass index (BMI) categories, normal (N), overweight (OW) and obese (OB), (D) metabolic syndrome, (E) kidney insufficiency, (F) current/past smokers (Yes/No), (G) former drinkers vs. lifetime abstainers, (H) current non-excessive drinkers vs. lifetime abstainers, (I) current excessive drinkers vs. lifetime abstainers, (J) poverty (Yes/No), (K) blood levels of lead and (L) cadmium (Q1-3 vs. Q4). Differences between groups were tested by univariate t statistic. Significance was a two sides P value less than 0.05, *P < 0.05, **P < 0.001, ***P < 0.0001. [a] Components of metabolic syndrome information was only available in participants with fasting blood test in NHANES 2007-2018, N=13,886. [b] Cadmium and lead analysis based on complete dataset with information of blood lead and cadmium measurements with N=42,255. Abbreviation: quartile, Q; standard error = SE.

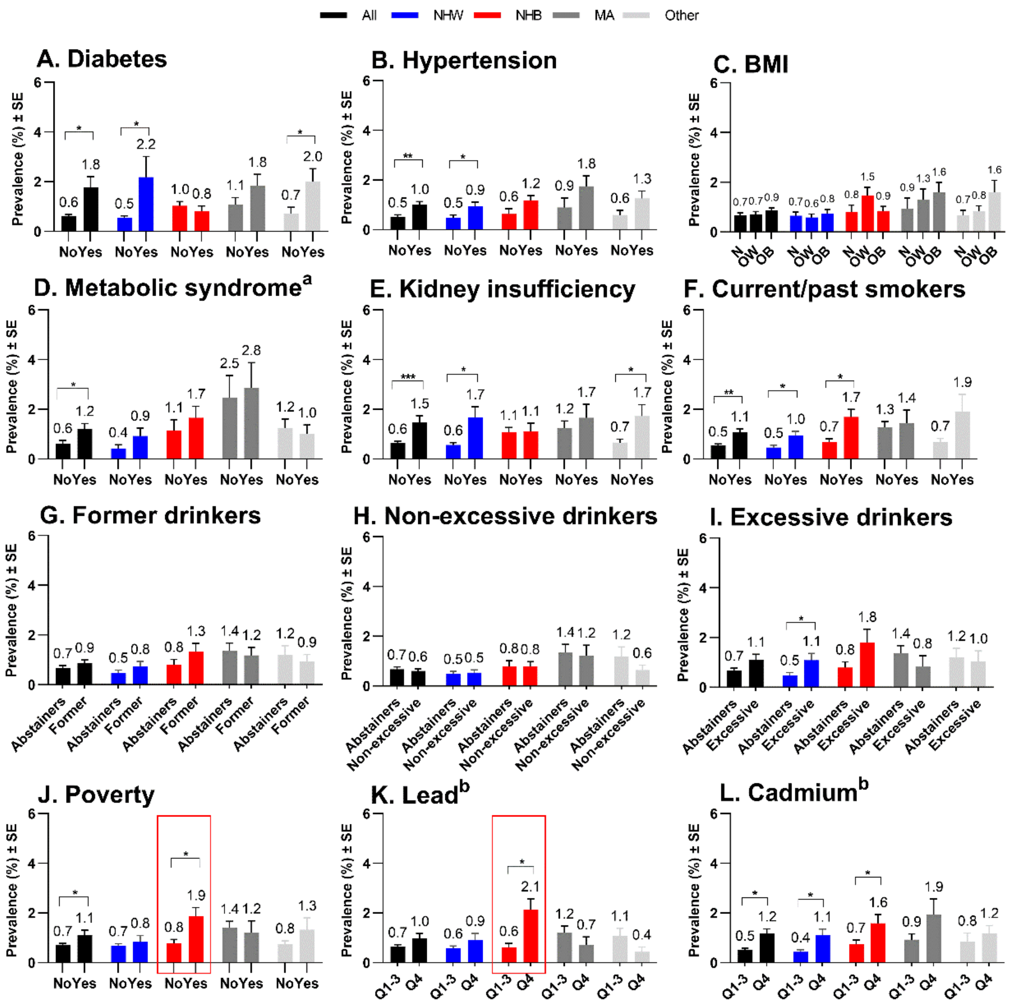

**Fig. S2. Age standardized weighted prevalence of advanced fibrosis in *females* aged 20-85 years, stratified by health conditions and race/ethnicity.** The age-standardized weighted prevalence of advanced fibrosis among female participants with and without various health conditions were determined for the total cohort (black), non-Hispanic White (NHW, blue), non-Hispanic Black (NHB, red), Mexican American (MA, grey) and other (light grey) racial/ethnic groups. (A) Diabetes (Yes/No), (B) hypertension (Yes/No), (C) body mass index (BMI) categories, normal (N), overweight (OW) and obese (OB), (D) metabolic syndrome, (E) kidney insufficiency, (F) current/past smokers (Yes/No), (G) former drinkers vs. lifetime abstainers, (H) current non-excessive drinkers vs. lifetime abstainers, (I) current excessive drinkers vs. lifetime abstainers, (J) poverty (Yes/No), (K) blood levels of lead and (L) cadmium (Q1-3 vs. Q4). Differences between groups were tested by univariate t statistic. Significance was a two sides P value less than 0.05, *P < 0.05, **P < 0.001, ***P < 0.0001. [a] Components of metabolic syndrome information was only available in participants with fasting blood test in NHANES 2007-2018, N=13,886. [b] Cadmium and lead analysis based on complete dataset with information of blood lead and cadmium measurements with N=42,255. Abbreviation: quartile, Q; standard error = SE.

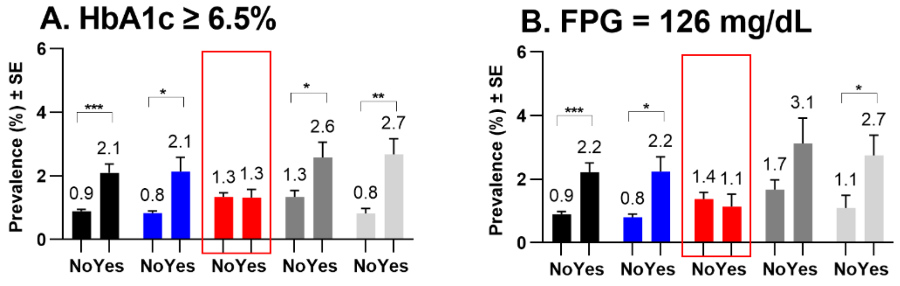

**Fig. S3. Age standardized weighted prevalence of advanced fibrosis, stratified by alternative definition of diabetes and race/ethnicity.** The age-standardized weighted prevalence of advanced fibrosis stratified by alternative definitions of diabetes including (A) hemoglobin A1c (HbA1c) >= 6.5%, (B) fasting plasma glucose (FPG) >= 126 mg/Dl in the total cohort (black), non-Hispanic White (NHW, blue), non-Hispanic Black (NHB, red), Mexican American (MA, grey) and other (light grey) racial/ethnic groups. Differences between groups were tested by univariate t statistic. Significance was a two sides P value less than 0.05, *P <0.05, **P <0.001, ***P <0.0001. Abbreviation: quartile, Q; standard error, SE.

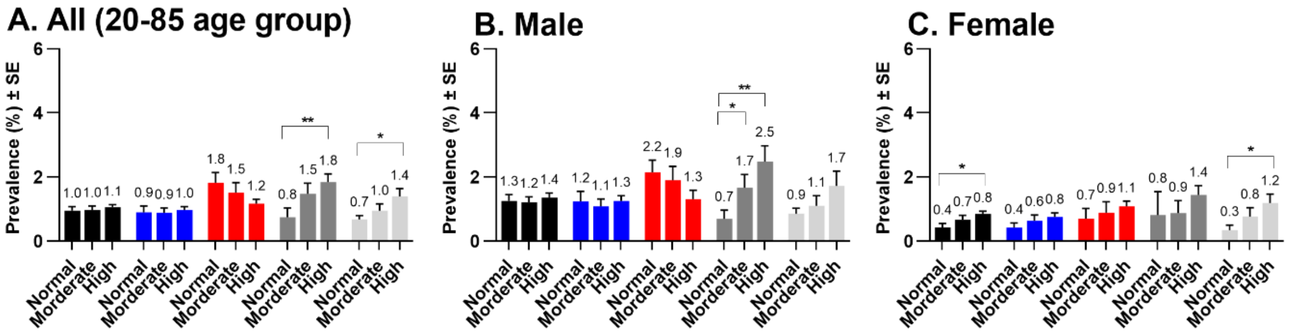

**Fig. S4. Age standardized weighted prevalence of advanced fibrosis, stratified by waist circumference and race/ethnicity.** The age-standardized weighted prevalence of advanced fibrosis stratified by sex and waist circumference among all (black), non-Hispanic White (NHW, blue), non-Hispanic Black (NHB, red), Mexican American (MA, grey) and other (light grey) racial/ethnic groups. waist circumstance categories, normal (N) (< 94 cm men, < 80 cm women), moderate (M) (>= 94 <102 cm men, >=80 < 88 cm women) and high (H) (>=102 cm men, >= 88 cm women). Differences between groups were tested by univariate t statistic. Significance was a two sides P value less than 0.05, *P <0.05, **P <0.001, ***P <0.0001. Abbreviation: quartile, Q; standard error, SE.

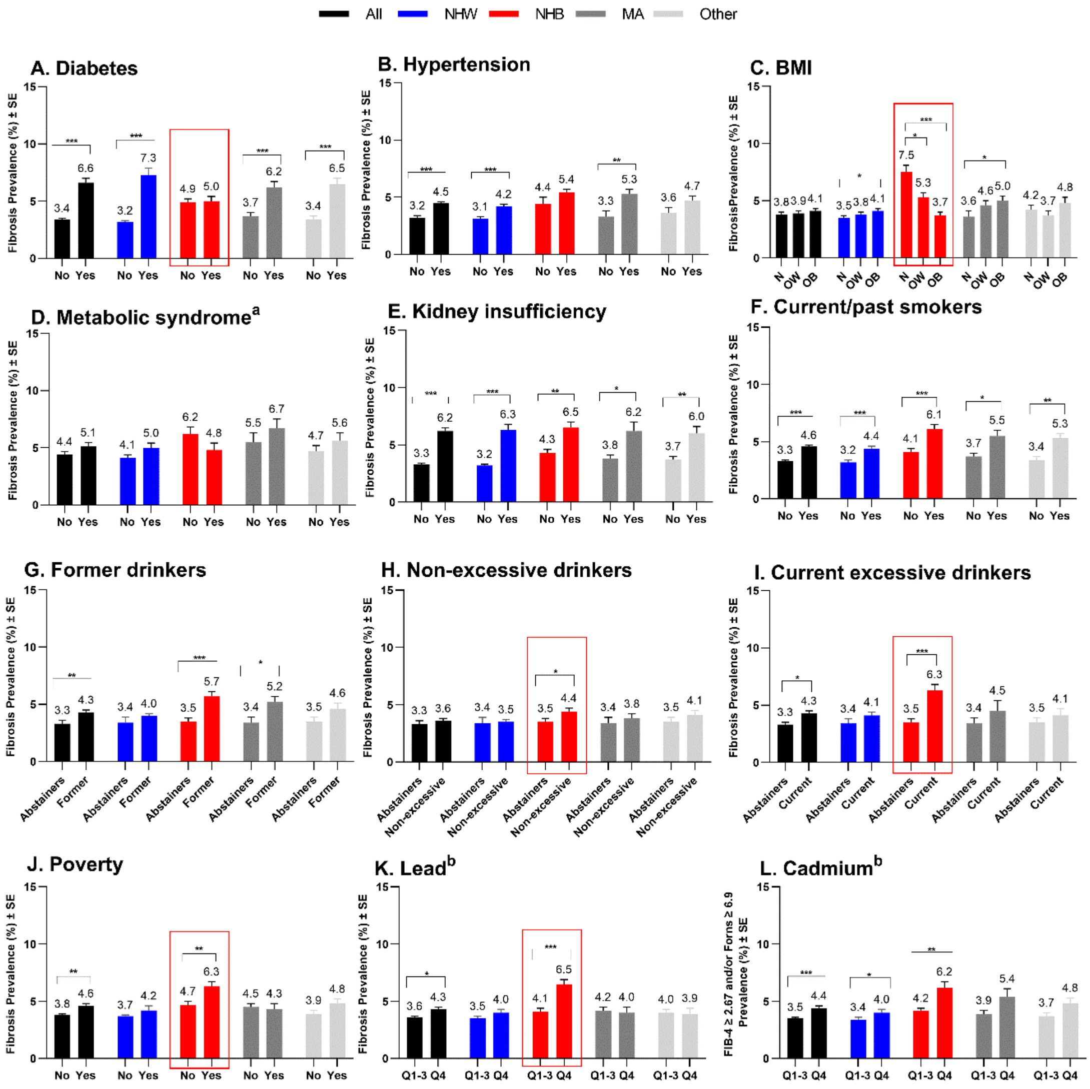

**Fig. S5. Age-standardized weighted prevalence of liver fibrosis (FIB-4 >= 2.67 and/or Forns >= 6.9) in participants aged 20-85 years, stratified by health conditions and race/ethnicity.** The age-standardized weighted prevalence of fibrosis (FIB-4 >= 2.67 and/or Forns >= 6.9) among participants with and without various health conditions were determined for the total cohort (black), non-Hispanic White (NHW, blue), non-Hispanic Black (NHB, red), Mexican American (MA, grey) and other (light grey) racial/ethnic groups. (A) Diabetes (Yes/No), (B) hypertension (Yes/No), (C) body mass index (BMI) categories, normal (N), overweight (OW) and obese (OB), (D) metabolic syndrome, (E) kidney insufficiency, (F) current/past smokers (Yes/No), (G) former drinkers vs. lifetime abstainers, (H) current non-excessive drinkers vs. lifetime abstainers, (I) current excessive drinkers vs. lifetime abstainers, (J) poverty (Yes/No), (K) blood levels of lead and (L) cadmium (Q1-3 vs. Q4). Differences between groups were tested by univariate t statistic. Significance was a two sides P value less than 0.05, *P < 0.05, **P < 0.001, ***P < 0.0001. [a] Components of metabolic syndrome information was only available in participants with fasting blood test in NHANES 2007-2018, N=13,886. [b] Cadmium and lead analysis based on complete dataset with information of blood lead and cadmium measurements with N=42,255. Abbreviation: quartile, Q; standard error = SE.

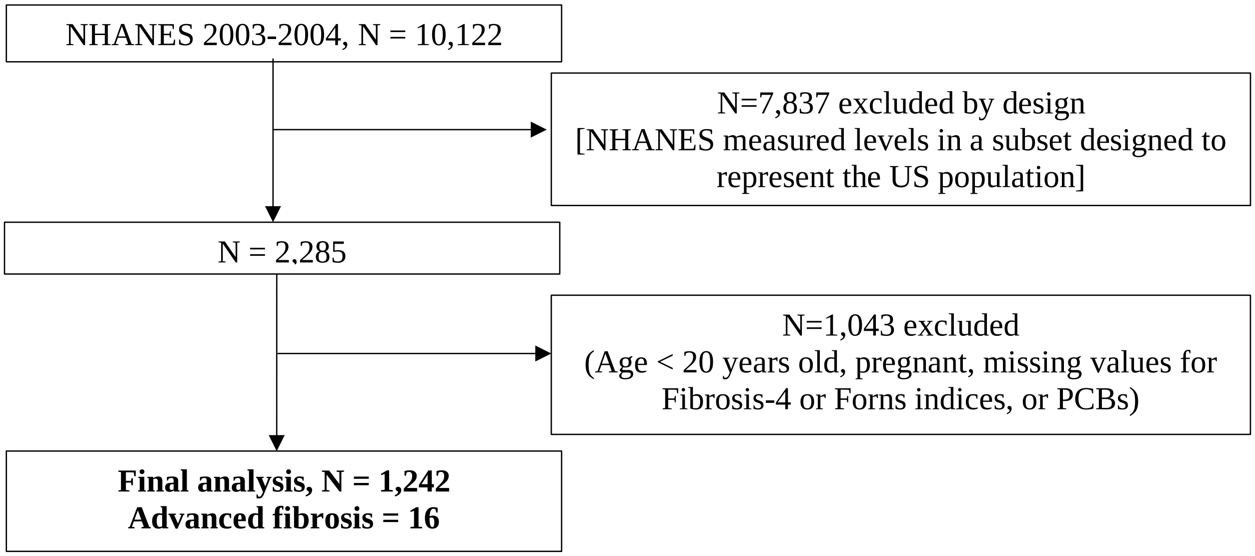

**Fig. S6. Flowchart showing the selection of participant in NHANES 2003-2004 for studies examining the prevalence of advanced fibrosis in people with data about polychlorinated biphenyl exposure.** Data from National Health and Nutrition Evaluation Survey (NHANES) 2003-2004 were used for the analysis of the relationship between advanced liver fibrosis and exposure to polychlorinated biphenyls (PCBs). Only compounds that were detected in at least 60% of lipid-adjusted serum measurements were included. Twenty-five non-dioxin-like PCBs and Nine coplanar PCBs (cPCBs) were included in the analysis. Total PCBs were the sum of the non-dioxin-like PCBs and the cPCBs. Four percentile values were used to categorize each PCBs. The cumulative measure of non-dioxin-like PCBs, cPCBs and total PCBs were derived from the sum of ranks of each PCB and were further categorized by four quartiles.

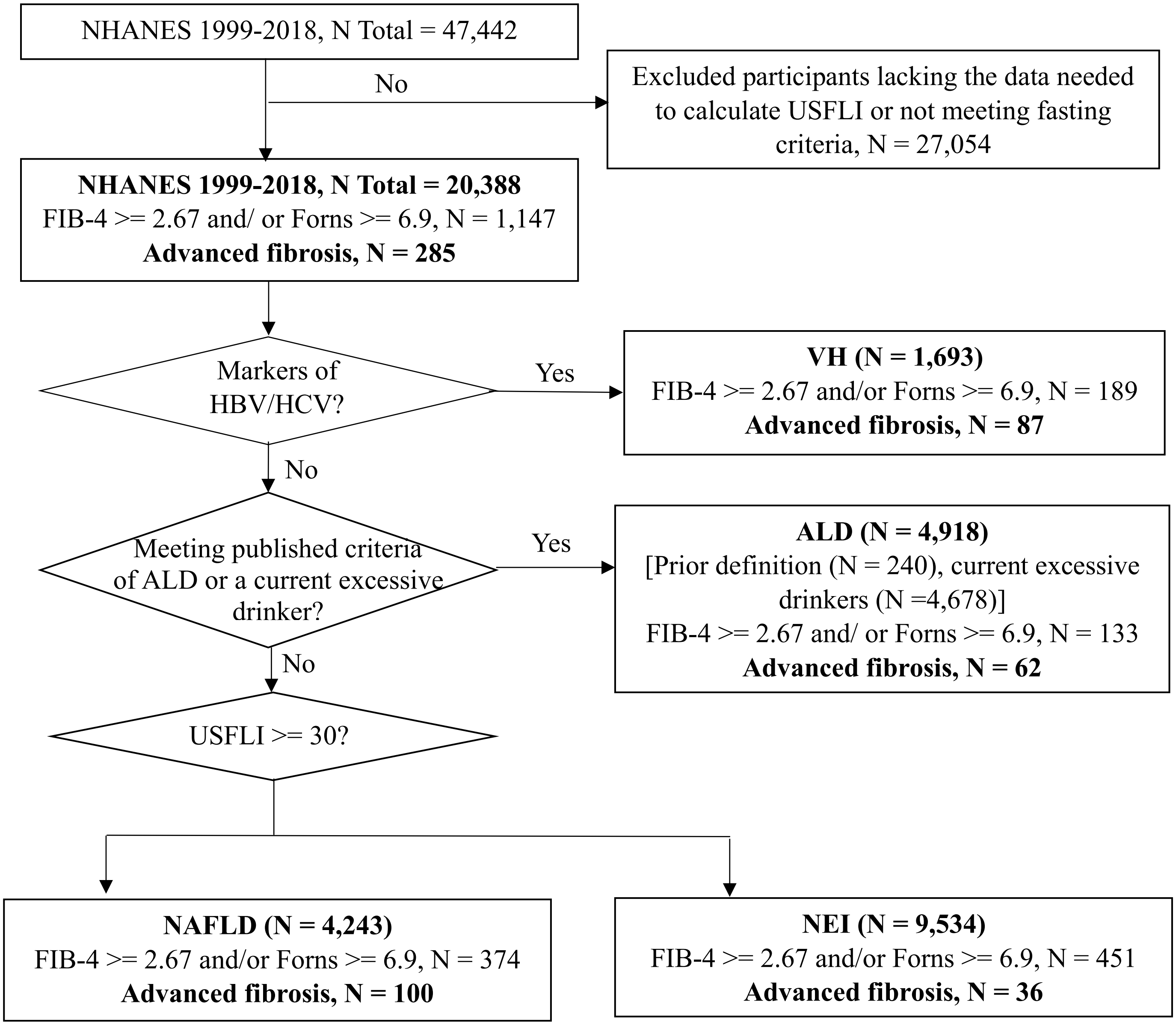

**Fig. S7. Flowchart of NHANES 1999-2018 showing eligibility for exposure analyses using USFLI to define NAFLD.** Liver disease exposures was analyzed among participants for whom the US Fatty Liver Index (USFLI) could be calculate included 20388 participants in National Health and Nutrition Evaluation Survey (NHANES) 1999-2018. Viral hepatitis (VH), past or current hepatitis B or hepatitis C virus infection; alcohol-associated liver disease (ALD), meeting previous criteria or current excessive alcohol use; non-alcoholic fatty liver disease (NAFLD), USFLI scores >= 30; No exposures identified (NEI), all others.

**A.**

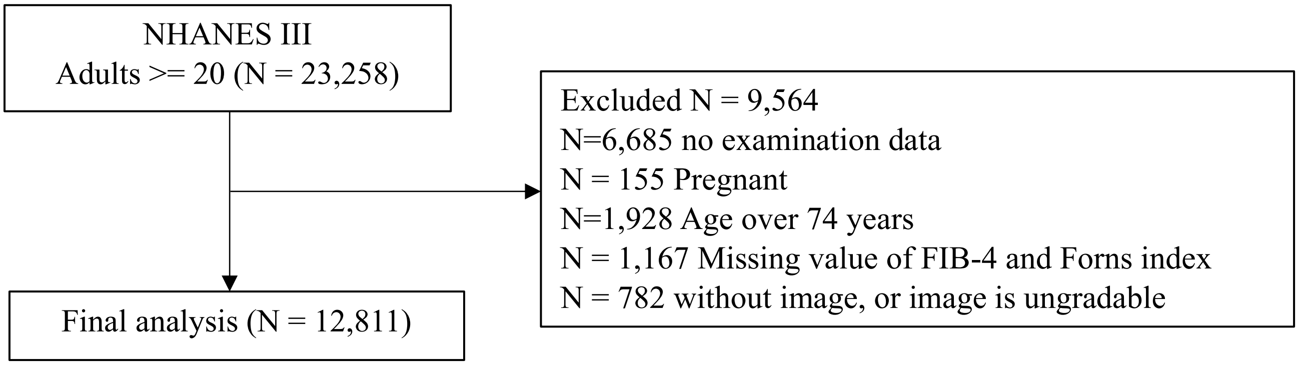

**B.**

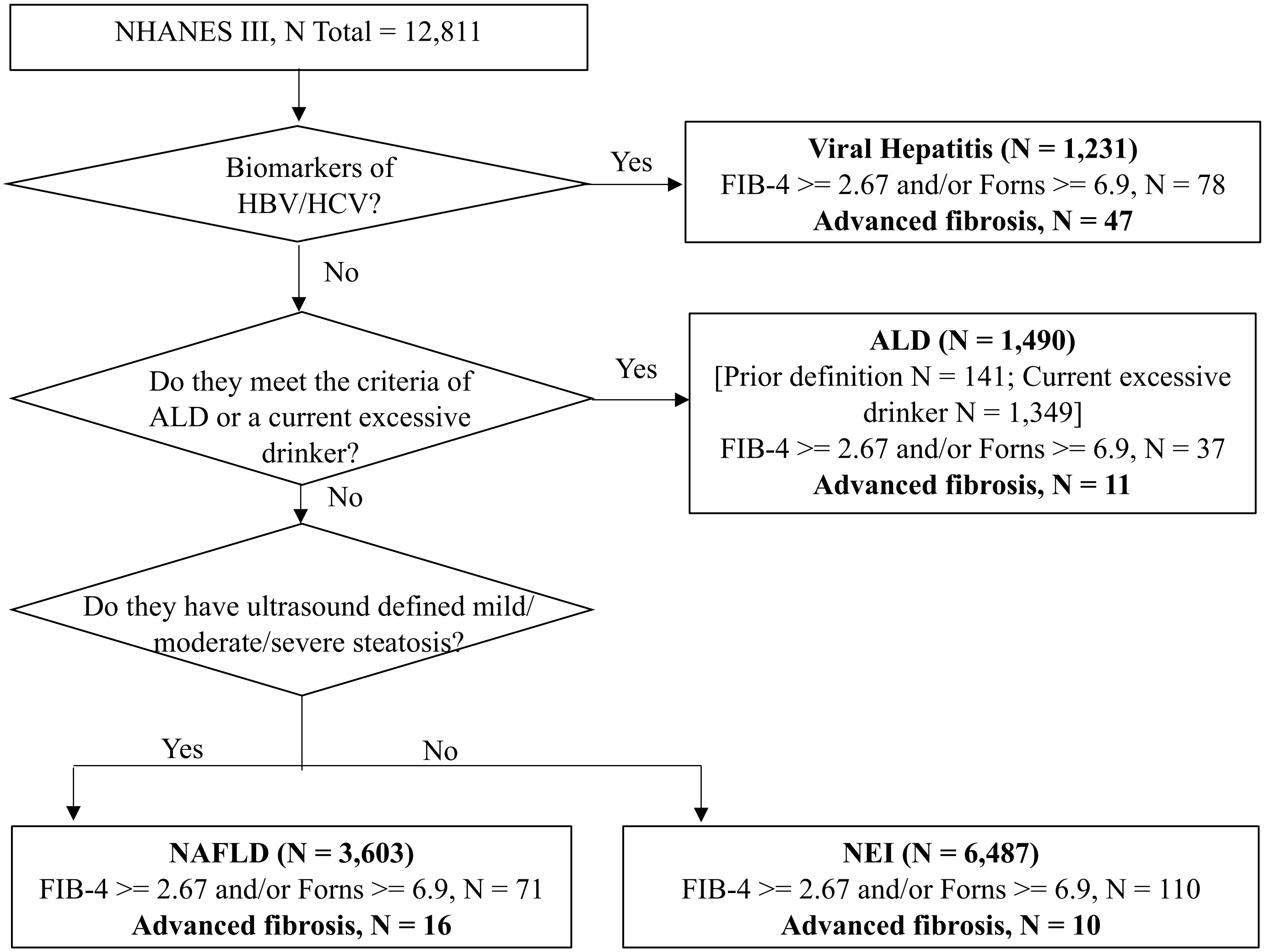

**Fig. S8. Flowchart of NHANES III (1988-1994) showing eligibility for exposure analyses using ultrasound to define NAFLD.** The study group included 12,811 participants in National Health and Nutrition Evaluation Survey (NHANES) III. A hierarchical method was used to assign participants to exposure groups: Viral hepatitis (VH), past or current hepatitis B or hepatitis C virus infection; alcohol-associated liver disease (ALD), meeting previous criteria or current excessive alcohol use; non-alcoholic fatty liver disease (NAFLD), liver ultrasound defined mild/moderate/severe fatty liver; no exposures identified (NEI), all others.

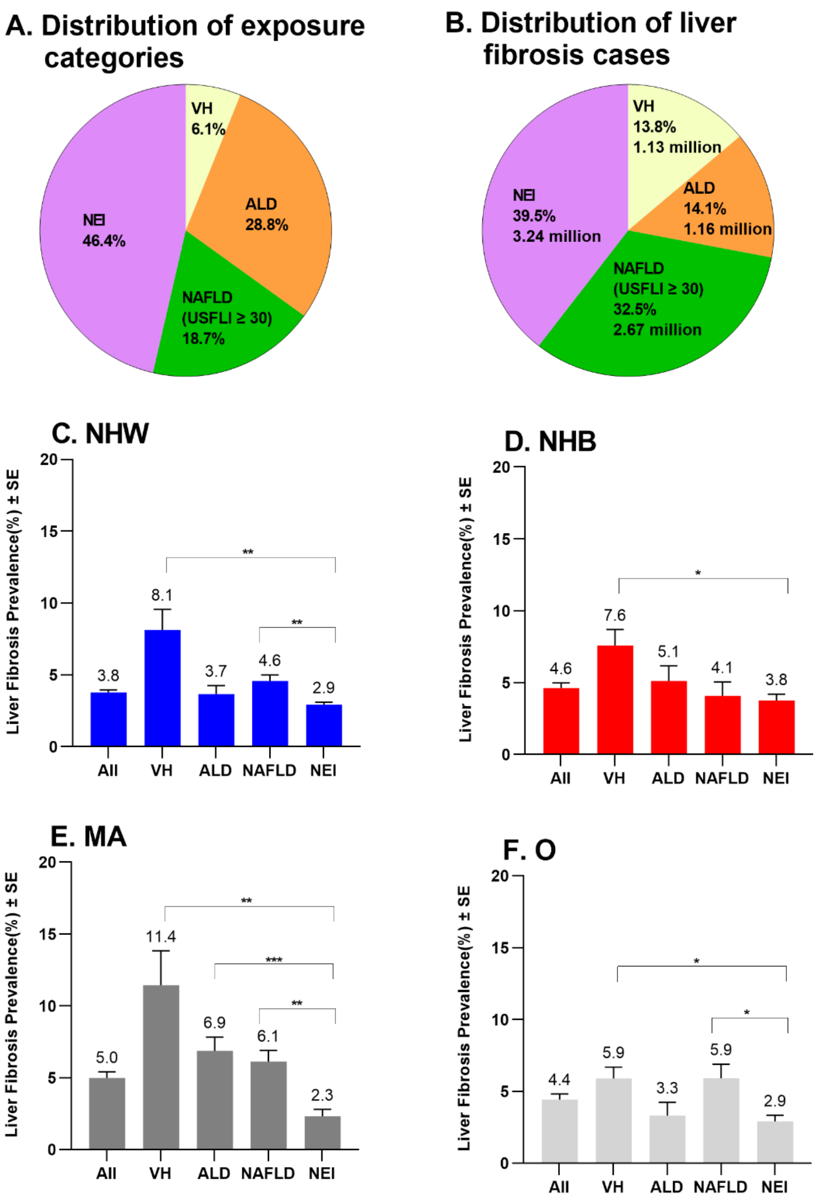

**Fig. S9. Liver fibrosis in people not meeting criteria for VH, ALD, or NAFLD**. Liver fibrosis defined by FIB-4 >= 2.67 and/or Forns >= 6.9. (A) the percentage of the total population meeting criteria for viral hepatitis (VH), alcohol-associated liver disease (ALD), or non-alcoholic fatty liver disease (NAFLD), and those with No exposure identified (NEI); (B) distribution of liver fibrosis cases in each category. The age-standardized weighted prevalence of liver fibrosis in each category among (C) non-Hispanic White (NHW), (D) non-Hispanic Black (NHB), (E) Mexican American (MA) and (F) other race (O). Differences between groups were tested by univariate t statistic, *P < 0.05, **P <0.001, ***P <0.0001. Abbreviation: standard error, SE.

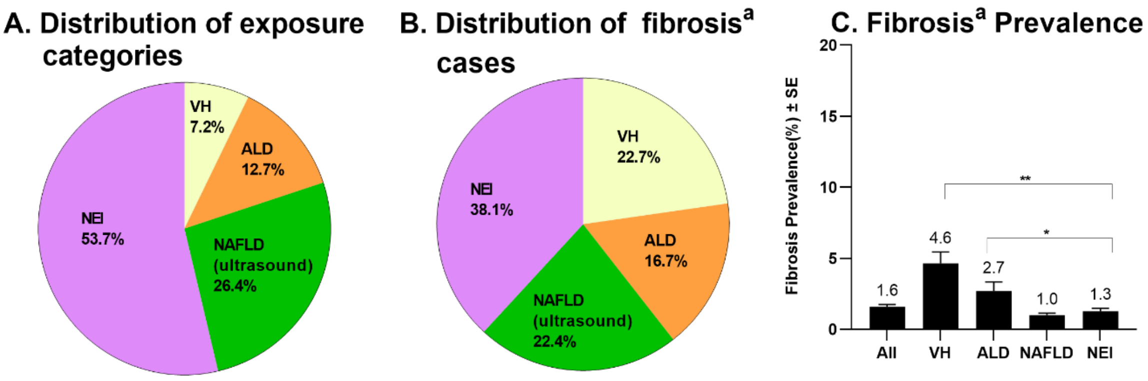

**Fig. S10. Liver fibrosis in people not meeting criteria for VH, ALD, or NAFLD in NHANES III population with ultrasound defined NAFLD**. The study group analyzed was based on participants in National Health and Nutrition Evaluation Survey (NHANES) III (1988-1994); in this group liver ultrasound data (normal vs. mild, moderate, and severe fatty liver disease) were used to define non-alcoholic fatty liver disease (NAFLD). (A) the percentage of the study group in each liver disease exposure category: viral hepatitis (VH), alcohol-associated liver disease (ALD), non-alcoholic fatty liver disease (NAFLD), and no exposures identified (NEI). (B) the percentage of the people with liver fibrosis in each exposure category. (C) the age-standardized weighted prevalence of advanced fibrosis in each exposure category. *P <0.05, **P <0.001. Data in NHANES III (1988-1994) was about 30 years earlier then NHANES 2017-2018. The prevalence of fibrosis in NHANES 2017-2018 [4.9% (95%CI, 4.2, 5.6), see table S1] was 3 times higher than NHANES III (1988-1994) [1.6% (95%CI, 1.3, 1.9), panel C] in total population. [a] Fibrosis defined by FIB-4 >= 2.67 and/or Forns >= 6.9.

**Table S1. Trends of advanced fibrosis and health conditions from 1999-2000 to 2017-2018**

| Variables | Group | Age-standardized weighted prevalence% (95% CI) | | Fold change[a] | Annual Percent Change [b] (95% CI) from 1999-2000 to 2017-2018 (ten cycles) | P trend |
|---|---|---|---|---|---|---|
| | | 1999-2000 | 2017-2018 | | | |
| **Advanced fibrosis** | **Total** | **0.7 (0.4, 1.0)** | **1.4 (1.2, 1.8)** | **2.0** | **8.7 (6.7, 10.9)** | **<0.001** |
| **FIB-4 ≥ 2.67 and/or Forns ≥ 6.9** | **Total** | **2.4 (2.0, 2.8)** | **4.9 (4.2, 5.6)** | **2.0** | **10.7 (6.7, 14.7)** | **<0.001** |
| **Kidney insufficiency** | **Total** | 13.9 (12.5, 15.3) | 13.5 (12.3, 14.8) | 0.97 | -0.1 (-1.4, 1.2) | 0.9 |
| | **NHW** | 12.4 (11.0, 13.8) | 12.1 (10.1, 14.0) | 0.98 | -0.1 (-1.8, 1.6) | 0.9 |
| | **NHB** | 20.5 (17.5, 23.4) | 20.2 (18.1, 22.4) | 0.99 | -0.0 (-1.2, 1.2) | 0.98 |
| | **MA** | 14.9 (11.8, 17.9) | 14.9 (13.5, 16.3) | 1.00 | 0.5 (-1.0, 1.9) | 0.5 |
| | **O** | 18.2 (14.8, 21.7) | 14.6 (12.9, 16.3) | 0.80 | -2.2 (-5.1, 0.7) | 0.1 |
| **Diabetes** | **Total** | **9.1 (8.0, 10.3)** | **14.7 (13.4, 16.0)** | **1.62** | **4.9 (3.9, 6.0)** | **<0.001** |
| | **NHW** | **7.7 (6.5, 9.0)** | **13.2 (11.4, 15.0)** | **1.71** | **5.4 (3.8, 7.1)** | **<0.001** |
| | **NHB** | 15.3 (13.4, 17.3) | 18.8 (16.5, 21.1) | 1.23 | 2.2 (-2.1, 6.6) | 0.3 |
| | **MA** | **12.5 (10.6, 14.4)** | **19.7 (17.2, 22.1)** | **1.58** | **4.5 (2.4, 6.7)** | **0.001** |
| | **O** | **12.0 (7.5, 16.5)** | **16.8 (15.3, 18.3)** | **1.40** | **2.1 (0.1, 4.1)** | **0.04** |
| **Hypertension** | **Total** | 48.3 (45.0, 51.5) | 48.6 (45.7, 51.4) | 1.01 | Segment1 (1999-2014): -0.1 (-2.1, 0.2)<br>Segment2 (2014-2018): 4.3 (-3.9, 13.1) | P1 = 0.08<br>P2 = 0.2 |
| | **NHW** | 47.3 (42.9, 51.7) | 47.2 (43.3, 51.2) | 1.00 | -0.4 (-1.1, 0.4) | 0.3 |
| | **NHB** | 58.6 (54.5, 62.6) | 60.5 (56.7, 64.3) | 1.03 | 0.1(-0.8, 0.9) | 0.9 |
| | **MA** | 44.8 (41.7, 47.9) | 46.7 (44.0, 49.3) | 1.04 | Segment1 (1999-2012): -1.9 (-4.8, 1.1)<br>Segment2 (2012-2018): 6.2 (-1.4, 14.4) | P1 =0 .2<br>P2 = 0.1 |
| | **O** | 47.4 (38.6, 56.2) | 48.5 (45.7, 51.4) | 1.02 | Segment1 (1999-2006): -10.0 (-24.2, 7.0)<br>**Segment2 (2006-2018): 3.7 (0.9, 6.5)** | P1 = 0.2<br>**P2 = 0.02** |
| **Obesity (BMI ≥ 30Kg/m$^2$)** | **Total** | **29.9 (26.8, 33.1)** | **42.8 (39.2, 46.5)** | **1.43** | **3.9 (3.1, 4.7)** | **<0.001** |
| | **NHW** | **28.0 (24.2, 31.9)** | **43.0 (37.6, 48.4)** | **1.54** | **4.0 (3.1, 5.0)** | **<0.001** |
| | **NHB** | **39.7 (36.2, 43.3)** | **50.6 (47.2, 53.9)** | **1.27** | **2.7 (1.6, 3.8)** | **<0.001** |
| | **MA** | **33.9 (28.2, 39.5)** | **50.4 (46.6, 54.3)** | **1.49** | **6.6 (4.9, 8.4)** | **<0.001** |
| | **O** | **28.7 (23.3, 34.2)** | **32.1 (28.3, 35.9)** | **1.12** | **2.8 (0.2, 5.5)** | **0.04** |
| | **Total** | **46.8 (42.2, 51.4)** | **60.2 (57.0, 63.5)** | **1.29** | **2.5 (1.9, 3.0)** | **<0.001** |
| | **NHW** | **46.2 (40.4, 52.0)** | **61.6 (57.1, 66.1)** | **1.33** | **2.6 (2.0, 3.1)** | **<0.001** |

| | | | | | | |
|---|---|---|---|---|---|---|
| **Obesity (WC ≥ 102cm for men, ≥ 88cm for women)** | **NHB** | **53.1 (50.1, 56.1)** | **62.9 (59.8, 66.1)** | **1.18** | **2.1 (1.3, 2.9)** | **<0.001** |
| | **MA** | **48.3 (44.4, 52.1)** | **68.0 (63.4, 72.5)** | **1.41** | **4.4 (3.7, 5.0)** | **<0.001** |
| | **O** | **44.3 (37.4, 51.1)** | **48.9 (45.2, 52.5)** | **1.10** | **1.4 (-0.0, 2.9)** | **0.05** |
| **Metabolic syndrome c** | **Total** | 36.9 (33.7, 40.0) | 38.2 (35.2, 41.2) | 1.04 | 1.2 (-1.5, 4.1) | 0.3 |
| | **NHW** | 38.0 (33.4, 42.6) | 37.5 (33.5, 41.4) | 0.99 | 1.4 (-2.2, 5.2) | 0.3 |
| | **NHB** | 34.3 (29.1, 39.5) | 35.9 (32.1, 39.7) | 1.04 | -0.9 (-4.6, 2.8) | 0.5 |
| | **MA** | 38.3 (34.2, 42.5) | 47.3 (42.9, 51.7) | 1.23 | 3.1 (-1.2, 7.5) | 0.1 |
| | **O** | 30.5 (23.8, 37.1) | 36.4 (32.3, 40.5) | 1.19 | 4.9 (-1.4, 11.6) | 0.1 |
| **Past/current smoking** | **Total** | **49.2 (45.9, 52.5)** | **42.5 (39.6, 45.4)** | **0.86** | Segment1 (1999-2004): 1.1 (-6.6, 9.6)<br>**Segment2 (2004-2018): -2.7 (-3.6, -1.8)** | P1 = 0.7<br>**P2 =0.001** |
| | **NHW** | **52.5 (48.5, 56.5)** | **45.9 (42.2, 49.6)** | **0.87** | **-2.0 (-2.6, -1.3)** | **<0.001** |
| | **NHB** | 42.9 (37.0, 48.7) | 38.4 (33.7, 43.2) | 0.90 | -1.2 (-2.5, 0.0) | 0.06 |
| | **MA** | **42.9 (38.7, 47.1)** | **37.2 (30.7, 43.6)** | **0.87** | **-3.6 (-5.2, -1.9)** | **0.001** |
| | **O** | 40.4 (34.8, 46.0) | 36.8 (33.4, 40.3) | 0.91 | -1.5 (-3.2, 0.3) | 0.09 |
| **Former drinker** | **Total** | **16.8 (14.8, 18.9)** | **16.5 (14.6, 18.4)** | **0.98** | **-2.7 (-4.7, -0.5)** | **0.02** |
| | **NHW** | **15.8 (13.2, 18.4)** | **15.4 (13.3, 17.5)** | **0.97** | **-2.3 (-4.4, -0.1)** | **0.04** |
| | **NHB** | **22.3 (17.5, 27.1)** | **18.3 (15.0, 21.7)** | **0.82** | **-4.0 (-7.3, -0.6)** | **0.03** |
| | **MA** | 19.6 (16.5, 22.6) | 16.4 (13.8, 19.0) | 0.84 | -1.8 (-3.8, 0.2) | 0.07 |
| | **O** | 18.5 (13.3, 23.6) | 19.9 (16.3, 23.6) | 1.08 | -1.0 (-5.3, 3.4) | 0.6 |
| **Non-current excessive drinker** | **Total** | 40.9 (38.3, 43.4) | 39.7 (37.5, 41.9) | 0.97 | -0.3 (-0.6, 0.0) | 0.08 |
| | **NHW** | 41.4 (38.1, 44.7) | 40.4 (36.8, 44.1) | 0.98 | -0.3 (-0.7, 0.2) | 0.2 |
| | **NHB** | 40.5 (36.2, 44.9) | 41.1 (37.4, 44.8) | 1.01 | 0.8 (-0.6, 2.1) | 0.2 |
| | **MA** | 33.9 (29.7, 38.1) | 34.6 (31.1, 38.0) | 1.02 | -0.4 (-1.8, 1.1) | 0.6 |
| | **O** | 40.9 (33.7, 48.0) | 37.4 (33.2, 41.5) | 0.91 | -0.3 (-1.6, 1.0) | 0.6 |
| **Current excessive drinker** | **Total** | **30.0 (28.4, 31.6)** | **36.8 (34.9, 38.6)** | **1.23** | **2.3 (1.5, 3.2)** | **<0.001** |
| | **NHW** | **32.2 (29.6, 34.7)** | **38.8 (36.0, 41.5)** | **1.20** | **2.4 (1.4, 3.5)** | **0.001** |
| | **NHB** | **18.3 (14.9, 21.6)** | **32.6 (29.3, 35.8)** | **1.78** | **5.8 (3.1, 8.5)** | **0.001** |
| | **MA** | 32.5 (29.9, 35.1) | 40.8 (36.5, 45.1) | 1.26 | 1.5 (-0.1, 3.0) | 0.06 |
| | **O** | 25.8 (21.1, 30.6) | 29.5 (25.0, 34.0) | 1.14 | 1.8 (-0.6, 4.3) | 0.1 |
| **Poverty** | **Total** | 13.7 (10.8, 16.6) | 12.1 (10.0, 14.1) | 0.88 | 1.5 (-2.4, 5.5) | 0.4 |
| | **NHW** | 8.9 (5.7, 12.2) | 8.6 (6.4, 10.9) | 0.97 | 0.6 (-3.5, 4.9) | 0.7 |
| | **NHB** | 24.3 (19.2, 29.3) | 20.9 (15.6, 26.2) | 0.86 | 1.7 (-2.0, 5.6) | 0.3 |

| | | | | | | |
|---|---|---|---|---|---|---|
| | **MA** | 26.6 (22.6, 30.6) | 19.7 (14.7, 24.7) | 0.74 | 1.2 (-2.8, 5.3) | 0.5 |
| | **O** | 28.3 (19.0, 37.6) | 17.1 (13.7, 20.6) | 0.60 | -1.8 (-6.5, 3.2) | 0.4 |
| **Q4 blood level of cadmium [d]** | **Total** | **28.0 (24.8, 31.2)** | **18.2 (16.4, 20.0)** | **0.65** | **-4.3 (-5.5, -3.1)** | **<0.001** |
| | **NHW** | **28.6 (24.4, 32.9)** | **17.8 (15.1, 20.6)** | **0.62** | **-4.8 (-6.3, -3.3)** | **<0.001** |
| | **NHB** | **30.6 (26.7, 34.4)** | **24.7 (21.4, 28.0)** | **0.81** | **-1.4 (-2.6, -0.2)** | **0.03** |
| | **MA** | **25.4 (20.4, 30.5)** | **10.0 (6.9, 13.1)** | **0.39** | **-7.7 (-10.4, -5.0)** | **<0.001** |
| | **O** | **25.1 (21.0, 29.3)** | **20.5 (17.6, 23.5)** | **0.82** | **-3.7 (-6.0, -1.2)** | **0.009** |
| **Q4 blood level of lead [d]** | **Total** | **35.0 (32.6, 37.3)** | **7.6 (6.2, 9.0)** | **0.22** | **-13.7 (-15.5, -11.9)** | **<0.001** |
| | **NHW** | **32.6 (30.2, 35.0)** | **6.5 (4.6, 8.3)** | **0.2** | **-13.7 (-15.4, -11.9)** | **<0.001** |
| | **NHB** | **41.6 (36.8, 46.4)** | **10.5 (8.7, 12.2)** | **0.25** | **-14.0 (-16.1, -11.7)** | **<0.001** |
| | **MA** | **43.7 (41.3, 46.0)** | **9.1 (4.9, 13.4)** | **0.21** | **-15.1 (-19.0, -11.0)** | **<0.001** |
| | **O** | **35.6 (29.8, 41.4)** | **8.6 (6.4, 10.9)** | **0.24** | **-14.3 (-11.10, -9.5)** | **<0.001** |

[a] Fold change calculated by the formula: Y / X; X was the age-standardized weighted prevalence of each condition in 1999-2000; Y was the age-standardized weighted prevalence of each condition in 2017-2018. [b] Annual Percent Change (APC) were measured trend in age-standardized weighted prevalence of advanced fibrosis and all other conditions from 1999-2000 to 2017-2018 based on Joinpoint analysis (prevalence data from 2001 to 2016 were not shown in tables). Significant increased/decreased trend colored in bold format.

[c] Components of metabolic syndrome information was only available in participants with fasting blood test in NHANES 2007-2018, N=13,886, the fold change calculated by the prevalence from 2017-2018 / 2007-2008. [d] Lead and cadmium analysis based on complete dataset with information of blood lead and cadmium measurements with N=42,255.

Abbreviations: alanine aminotransferase, ALT; confidence interval: CI; quartile: Q.

**Table S2. Odds ratios from multivariable logistic regression models with the outcome of advanced liver fibrosis in cohort excluded participants with viral hepatitis**

| **Cohort** | **In aged 20-85 years after excluded viral hepatitis, OR (95% CI)** | | | | |
|---|---|---|---|---|---|
| **Sample size (N)** | **Total (43,298)** | **NHW (20,203)** | **NHB (8,142)** | **MA (7,990)** | **O (6,963)** |
| **Advanced fibrosis cases (N)** | **415** | **196** | **60** | **97** | **62** |
| **Age (unit 10 years)** | **1.54 (1.41, 1.68) ‡** | **1.55 (1.36, 1.77) ‡** | **1.45 (1.21, 1.75) ‡** | **1.59 (1.32, 1.91) ‡** | **1.92 (1.55, 2.37) ‡** |
| **Kidney insufficiency** | | | | | |
| No | Reference | | | | |
| Yes | **1.40 (1.01, 1.95) *** | 1.53 (0.97, 2.41) | 1.05 (0.47, 2.35) | 1.17 (0.63, 2.16) | 1.11 (0.53, 2.34) |
| **Diabetes** | | | | | |
| No | Reference | | | | |
| Yes | **2.33 (1.76, 3.09) ‡** | **2.50 (1.75, 3.55) ‡** | 0.76 (0.39, 1.48) | 1.34 (0.72, 2.53) | **4.21 (2.14, 8.28) ‡** |
| **Hypertension** | | | | | |
| No | Reference | | | | |
| Yes | 1.33 (0.94, 1.89) | 1.36 (0.86, 2.16) | 0.64 (0.33, 1.26) | **3.37 (1.54, 7.40) *** | 0.94 (0.43, 2.06) |
| **BMI** | | | | | |
| Normal | Reference | | | | |
| Overweight | 1.02 (0.71, 1.46) | 1.02 (0.64, 1.63) | 1.08 (0.53, 2.20) | 1.44 (0.57, 3.63) | 0.58 (0.24, 1.36) |
| Obese | 1.18 (0.80, 1.74) | 1.21 (0.75, 1.97) | 0.81 (0.39, 1.67) | 1.29 (0.54, 3.06) | 1.32 (0.52, 3.40) |
| **Smoking status** | | | | | |
| Never | Reference | | | | |
| Past/current | **1.60 (1.21, 2.10) †** | **1.55 (1.07, 2.24) *** | **3.00 (1.56, 5.78) †** | 1.35 (0.80, 2.29) | 1.64 (0.84, 3.23) |

Multiple imputation was performed in univariate and multivariate logistic regression models. Variables with P value < 0.1 in the univariate in total cohort were included into multivariate analysis. *P <0.05, †P <0.001, ‡P <0.0001. Abbreviations: odds ratio, OR; confidence interval, CI; body mass index, BMI; Non-Hispanic White, NHW; Non-Hispanic Black, NHB; Mexican American, MA; other race, O.

**Table S3. Odds ratios from multivariable logistic regression models with the outcome of advanced liver fibrosis in people aged 20-85 years in NHANES 2007-2018 with metabolic syndrome included as a variable rather than diabetes, hypertension, or obesity**

| **Sample size (N)** | **Total (13,886)** |
|---|---|
| **Advanced fibrosis cases (N)** | **N= 227** |
| **Age (unit 10 years)** | **1.63 (1.47, 1.81) ‡** |
| **Gender** | |
| Female | Reference |
| Male | **1.75 (1.13, 2.69) *** |
| **Kidney insufficiency** | |
| No | Reference |
| Yes | 1.43 (0.91, 2.25) |
| **Metabolic syndrome [a]** | |
| No | Reference |
| Yes | **1.48 (1.03, 2.11) *** |
| **Alcohol use** | |
| Lifetime abstainers | Reference |
| Former drinkers | 0.68 (0.31, 1.47) |
| Non-excessive current drinkers | 0.62 (0.30, 1.29) |
| Excessive current drinkers | 1.40 (0.65, 3.05) |
| **Smoking status** | |
| Never | Reference |
| Past/current | **1.66 (1.12, 2.46) *** |
| **Poverty** | |
| No | Reference |
| Yes | **1.64 (1.05, 2.56) *** |

Multiple imputation was performed in multivariate logistic regression. Variables with P value < 0.1 in the univariate in total cohort were included into multivariate analysis. *P < 0.05, ‡P <0.0001. Components of metabolic syndrome information was only available in participants with fasting blood test in NHANES 2007-2018, N=13,886. Abbreviations: alanine aminotransferase: ALT; odds ratio, OR; confidence interval, CI.

**Table S4. Odds ratios from multivariable logistic regression models with the outcome of advanced liver fibrosis in aged 35-64 cohort**

| **Cohort** | **Aged 35-64 cohort, OR (95% CI)** | | | | |
|---|---|---|---|---|---|
| **Sample size (N)** | **Total (2,4001)** | **NHW (9,812)** | **NHB (5,201)** | **MA (4,447)** | **O (4,541)** |
| **Advanced fibrosis cases (N)** | **349** | **108** | **100** | **78** | **63** |
| **Age (unit 10 years)** | **1.92 (1.63, 2.35) ‡** | **1.90 (1.48, 2.45) ‡** | **1.65 (1.29, 2.11) ‡** | **1.87 (1.24, 2.82) *** | **3.50 (2.17, 5.64) ‡** |
| **Gender** | | | | | |
| Female | Reference | | | | |
| Male | **1.89 (1.30, 2.74) †** | **1.94 (1.10, 3.42) *** | **1.65 (1.03, 2.63) *** | 1.28 (0.58, 2.80) | **2.39 (1.19, 4.81) *** |
| **Kidney insufficiency** | | | | | |
| No | Reference | | | | |
| Yes | **1.77 (1.20, 2.60) *** | **2.12 (1.15, 3.89) *** | 1.34 (0.82, 2.20) | 0.93 (0.33, 2.69) | 1.67 (0.80, 3.41) |
| **Diabetes** | | | | | |
| No | Reference | | | | |
| Yes | **2.09 (1.50, 2.92) ‡** | **3.09 (1.92, 4.95) ‡** | **0.56 (0.32, 0.96) *** | 1.43 (0.64, 3.17) | **1.91 (1.00, 3.67) *** |
| **Hypertension** | | | | | |
| No | Reference | | | | |
| Yes | **1.51 (1.06, 2.17) *** | 1.55 (0.91, 2.63) | 1.02 (0.63, 1.65) | **2.47 (1.33, 4.56) *** | 1.23 (0.54, 2.77) |
| **BMI** | | | | | |
| Normal | Reference | | | | |
| Overweight | 0.69 (0.39, 1.14) | 0.68 (0.32, 1.43) | 0.70 (0.37, 1.33) | 0.84 (0.34, 2.01) | 0.58 (0.28, 1.21) |
| Obese | 0.63 (0.39, 1.01) | **0.48 (0.23, 0.98) *** | 0.56 (0.29, 1.08) | 0.84 (0.34, 2.09) | 1.70 (0.81, 3.57) |
| **Alcohol use** | | | | | |
| Lifetime abstainers | Reference | | | | |
| Former drinkers | 1.20 (0.57, 2.51) | 1.09 (0.24, 4.85) | 2.23 (0.97, 5.12) | 1.32 (0.57, 3.10) | 1.12 (0.46, 2.69) |
| Non-excessive current drinkers | 1.11 (0.54, 2.27) | 1.55 (0.28, 5.36) | 1.68 (0.71, 4.00) | 1.19 (0.56, 2.50) | 0.60 (0.23, 1.55) |
| Excessive current drinkers | **2.86 (1.39, 5.90) *** | 3.12 (0.73, 13.37) | **2.91 (1.34, 6.31) *** | **3.82 (1.30, 11.19) *** | 2.05 (0.81, 5.16) |
| **Smoking status** | | | | | |

| | | | | | |
|---|---|---|---|---|---|
| Never | Reference | | | | |
| Past/current | **1.73 (1.24, 2.43) *** | **1.76 (1.03, 3.02) *** | **1.82 (1.09, 3.03) *** | 1.97 (0.94, 4.13) | 1.39 (0.70, 2.77) |
| **Poverty** | | | | | |
| No | Reference | | | | |
| Yes | **1.88 (1.34, 2.65) †** | **1.82 (1.02, 3.25) *** | **2.47 (1.64, 3.73) ‡** | 1.64 (0.86, 3.13) | 0.77 (0.42, 1.41) |

Multiple imputation was performed in multivariate logistic regression models. Variables with P value < 0.1 in the univariate in total cohort were included into multivariate analysis. *P <0.05, †P <0.001, ‡P <0.0001.

Abbreviations: odds ratio, OR; confidence interval, CI; body mass index, BMI; Non-Hispanic White, NHW; Non-Hispanic Black, NHB; Mexican American, MA; other race, O.

**Table S5. Odds ratios from multivariable logistic regression models with the outcome of fibrosis defined by FIB-4 >= 2.67 and/or Forns >= 6.9**

| **Cohort** | **Fibrosis defined by FIB-4 ≥ 2.67 and/or Forns ≥ 6.9 in 20-85 years cohort, OR (95% CI)** | | | | |
|---|---|---|---|---|---|
| **Sample size (N)** | **Total (47,442)** | **NHW (21,167)** | **NHB (9,634)** | **MA (8,334)** | **Other (8,307)** |
| **Fibrosis cases (N)** | **2,745** | **1,480** | **559** | **338** | **368** |
| **Age (unit 10 years)** | **2.75 (2.60-2.90) ‡** | **2.85 (2.63-3.08) ‡** | **2.44 (2.23-2.67) ‡** | **2.52 (2.17-2.92) ‡** | **3.20 (2.86-3.58) ‡** |
| **Gender** | | | | | |
| Female | Reference | | | | |
| Male | **2.25 (2.01-2.52) ‡** | **2.28 (1.97-2.63) ‡** | **2.11 (1.65-2.69) ‡** | **1.76 (1.24-2.49) *** | **2.07 (1.47-2.93) ‡** |
| **Kidney insufficiency** | | | | | |
| No | Reference | | | | |
| Yes | **1.52 (1.32-1.75) ‡** | **1.54 (1.30-1.82) ‡** | **1.35 (1.05-1.73) *** | **1.52 (1.02-2.25) *** | **1.37 (1.00-1.86) *** |
| **Diabetes** | | | | | |
| No | Reference | | | | |
| Yes | **1.65 (1.43-1.90) ‡** | **1.79 (1.49-2.14) ‡** | 0.99 (0.79-1.23) | 1.39 (0.98-1.96) | **1.67 (1.18-2.37) *** |
| **Hypertension** | | | | | |
| No | Reference | | | | |
| Yes | **1.24 (1.07-1.43) *** | **1.18 (1.00-1.40) *** | 1.24 (0.92-1.67) | **1.66 (1.14-2.42) *** | 1.29 (0.86-1.94) |
| **BMI** | | | | | |
| Normal | Reference | | | | |
| Overweight | 0.85 (0.72-1.01) | 0.90 (0.73-1.11) | **0.68 (0.53-0.89) *** | 1.22 (0.80-1.86) | **0.67 (0.46-0.98) *** |
| Obese | 0.87 (0.75-1.00) | 0.91 (0.76-1.09) | **0.52 (0.39-0.68) †** | 1.25 (0.82-1.88) | 1.07 (0.69-1.66) |
| **Alcohol use** | | | | | |
| Lifetime abstainers | Reference | | | | |
| Former drinkers | 1.05 (0.85-1.30) | 1.00 (0.76-1.31) | 1.35 (0.94-1.95) | 1.38 (0.79-2.42) | 1.13 (0.76-1.66) |
| Non-excessive current drinkers | 0.99 (0.81-1.22) | 0.98 (0.75-1.27) | 1.17 (0.80-1.70) | 0.93 (0.54-1.60) | 1.01 (0.69-1.48) |
| Excessive current drinkers | **1.45 (1.13-1.86) *** | **1.38 (1.01-1.88) *** | **1.87 (1.25-2.84) *** | 1.93 (0.98-3.84) | 1.49 (0.92-2.42) |
| **Smoking status** | | | | | |
| Never | Reference | | | | |

| | | | | | |
|---|---|---|---|---|---|
| Past/current | **1.15 (1.00-1.32) *** | 1.13 (0.94-1.35) | 1.17 (0.91-1.49) | 1.24 (0.87-1.75) | 1.31 (0.97-1.75) |
| **Poverty** | | | | | |
| No | Reference | | | | |
| Yes | **1.29 (1.11-1.49)** † | 1.17 (0.93-1.48) | **1.46 (1.16-1.84)** † | 1.14 (0.80-1.63) | 1.17 (0.84-1.63) |

Multiple imputation was performed in multivariate logistic regression models. Variables with P value < 0.1 in the univariate in total cohort were included into multivariate analysis. *P <0.05, †P <0.001, ‡P <0.0001.

Abbreviations: odds ratio, OR; confidence interval, CI; body mass index, BMI; Non-Hispanic White, NHW; Non-Hispanic Black, NHB; Mexican American, MA; other race, O.

**Table S6. Odds ratios from multivariable logistic regression models with the outcome of advanced liver fibrosis and added blood cadmium level as continuous variable into models**

| **Cohort** | **In aged 20-85 years with cadmium into models, OR (95% CI)** | | | | |
|---|---|---|---|---|---|
| **Sample size (N)** | **Total (42,255)** | **NHW (19,176)** | **NHB (8,585)** | **MA (7,508)** | **O (6,986)** |
| **Advanced fibrosis cases (N)** | **542** | **226** | **128** | **102** | **86** |
| **Survey year** | **1.07 (1.03, 1.11) ‡** | **1.08 (1.03, 1.13) †** | 1.01 (0.95, 1.08) | 1.06 (0.97, 1.14) | 1.00 (0.90, 1.11) |
| **Age (unit 10 years)** | **1.57 (1.45, 1.69) ‡** | **1.52 (1.36, 1.70) ‡** | **1.54 (1.33, 1.78) ‡** | **1.81 (1.47, 2.22) ‡** | **2.11 (1.78, 2.51) ‡** |
| **Gender** | | | | | |
| Female | Reference | | | | |
| Male | **1.59 (1.23, 2.06) †** | **1.67 (1.15, 2.42) *** | **1.66 (1.13, 2.44) *** | 1.24 (0.68, 2.27) | 1.19 (0.63, 2.22) |
| **Kidney insufficiency** | | | | | |
| No | Reference | | | | |
| Yes | 1.17 (0.86, 1.58) | 1.18 (0.76, 1.82) | 1.16 (0.74, 1.81) | 0.61 (0.37, 1.01) | 1.55 (0.84, 2.87) |
| **Diabetes** | | | | | |
| No | Reference | | | | |
| Yes | **2.34 (1.79, 3.06) ‡** | **2.81 (2.03, 3.91) ‡** | 0.75 (0.49, 1.16) | **1.89 (1.01, 3.55)** | **2.55 (1.26, 5.17) *** |
| **Hypertension** | | | | | |
| No | Reference | | | | |
| Yes | **1.86 (1.34, 2.58) †** | **2.03 (1.27, 3.25) *** | 1.34 (0.78, 2.29) | **2.36 (1.14, 4.87) *** | 1.12 (0.54, 2.34) |
| **BMI** | | | | | |
| Normal | Reference | | | | |
| Overweight | 0.88 (0.60, 1.30) | 0.85 (0.49, 1.47) | 0.89 (0.51, 1.57) | 1.10 (0.53, 2.28) | 0.89 (0.41, 1.94) |
| Obese | 0.88 (0.63, 1.24) | 0.79 (0.49, 1.26) | 0.78 (0.44, 1.38) | 0.97 (0.43, 2.16) | 1.81 (0.74, 4.42) |
| **Alcohol use** | | | | | |
| Lifetime abstainers | Reference | | | | |
| Former drinkers | 0.92 (0.61, 1.39) | 0.87 (0.50, 1.50) | 1.94 (0.87, 4.33) | 0.83 (0.30, 2.30) | 0.85 (0.34, 2.16) |
| Non-excessive current drinkers | 0.99 (0.66, 1.51) | 1.08 (0.62, 1.87) | 1.44 (0.60, 3.44) | 0.76 (0.25, 2.36) | 0.74 (0.31, 1.77) |
| Excessive current drinkers | **2.01 (1.29, 3.12) *** | **2.09 (1.13, 3.87) *** | **2.59 (1.15, 5.80) *** | 2.00 (0.63, 6.32) | 1.68 (0.64, 4.37) |

| | | | | | |
|---|---|---|---|---|---|
| **Smoking status** | | | | | |
| Never | Reference | | | | |
| Past/current | **1.50 (1.13, 1.99) *** | **1.49 (1.02, 2.18) *** | 1.20 (0.76, 1.92) | 1.62 (0.83, 3.14) | 1.87 (0.85, 4.11) |
| **Poverty** | | | | | |
| No | Reference | | | | |
| Yes | **1.32 (1.00, 1.76) *** | 1.03 (0.64, 1.66) | **1.91 (1.23, 2.96) *** | 1.17 (0.61, 2.24) | 1.07 (0.57, 2.02) |
| **Blood level of cadmium (μg/L)** | **1.34 (1.22, 1.48) ‡** | **1.35 (1.19, 1.53) ‡** | **1.70 (1.38, 2.08) ‡** | 1.23 (0.96, 1.59) | 1.14 (0.88, 1.48) |

Cadmium analysis based on complete dataset with information of blood lead and cadmium measurements with N=42,255. Multiple imputation was performed in univariate and multivariate logistic regression models. Variables with P value < 0.1 in the univariate in total cohort were included into multivariate analysis. *P < 0.05, †P < 0.001, ‡P <0.0001. Abbreviations: odds ratio, OR; confidence interval, CI; body mass index, BMI; Non-Hispanic White, NHW; Non-Hispanic Black, NHB; Mexican American, MA; other race, O.

**Table S7. Odds ratios from multivariable logistic regression models with the outcome of advanced liver fibrosis and added blood lead level as continuous variable into models**

| **Cohorts** | **In aged 20-85 years with lead into models, OR (95% CI)** | | | | |
|---|---|---|---|---|---|
| **Sample size (N)** | **Total (42,255)** | **NHW (19,176)** | **NHB (8,585)** | **MA (7,508)** | **O (6,986)** |
| **Advanced fibrosis cases (N)** | **542** | **226** | **128** | **102** | **86** |
| **Survey year** | **1.07 (1.04, 1.11) ‡** | **1.09 (1.04, 1.14) †** | 1.03 (0.97, 1.10) | 1.07 (0.98, 1.16) | 1.00 (0.91, 1.12) |
| **Age (unit 10 years)** | **1.53 (1.42, 1.65) ‡** | **1.48 (1.32, 1.65) ‡** | **1.46 (1.26, 1.69) ‡** | **1.83 (1.48, 2.26) ‡** | **2.15 (1.80, 2.58) ‡** |
| **Gender** | | | | | |
| Female | Reference | | | | |
| Male | **1.47 (1.13, 1.92) †** | **1.54 (1.05, 2.25) *** | 1.41 (0.94, 2.12) | 1.24 (0.69, 2.24) | 1.24 (0.7, 2.21) |
| **Kidney insufficiency** | | | | | |
| No | Reference | | | | |
| Yes | 1.19 (0.88, 1.61) | 1.21 (0.79, 1.86) | 1.16 (0.73, 1.84) | 0.64 (0.38, 1.06) | 1.60 (0.86, 2.96) |
| **Diabetes** | | | | | |
| No | Reference | | | | |
| Yes | **2.36 (1.81, 3.07) ‡** | **2.84 (2.04, 3.95) ‡** | 0.79 (0.51, 1.20) | 1.84 (0.99, 3.43) | **2.53 (1.26, 5.07) *** |
| **Hypertension** | | | | | |
| No | Reference | | | | |
| Yes | **1.87 (1.34, 2.60) †** | **2.04 (1.27, 3.28) *** | 1.30 (0.76, 2.23) | **2.37 (1.15, 4.89) *** | 1.12 (0.53, 2.36) |
| **BMI** | | | | | |
| Normal | Reference | | | | |
| Overweight | 0.85 (0.58, 1.25) | 0.82 (0.47, 1.42) | 0.88 (0.50, 1.53) | 1.07 (0.51, 2.26) | 0.87 (0.4, 1.85) |
| Obese | 0.85 (0.60, 1.19) | 0.76 (0.48, 1.21) | 0.75 (0.42, 1.33) | 0.93 (0.41, 2.09) | 1.70 (0.70, 4.12) |
| **Alcohol use** | | | | | |
| Lifetime abstainers | Reference | | | | |
| Former drinkers | 0.91 (0.61, 1.37) | 0.86 (0.50, 1.49) | 1.77 (0.79, 3.95) | 0.83 (0.30, 2.29) | 0.86 (0.35, 2.13) |
| Non-excessive current drinkers | 0.97 (0.64, 1.47) | 1.04 (0.60, 1.80) | 1.40 (0.59, 3.32) | 0.79 (0.26, 2.42) | 0.76 (0.32, 1.79) |
| Excessive current drinkers | 0.91 (0.61, 1.37) | **2.06 (1.12, 3.77) *** | **2.64 (1.18, 5.87) *** | 2.07 (0.66, 6.56) | 1.75 (0.69, 4.49) |

| | | | | | |
|---|---|---|---|---|---|
| **Smoking status** | | | | | |
| Never | Reference | | | | |
| Past/current | **1.70 (1.29, 2.24) *** | **1.68 (1.15, 2.45) †** | **1.58 (1.03, 2.43) *** | 1.76 (0.93, 3.33) | 1.97 (0.91, 4.25) |
| **Poverty** | | | | | |
| No | Reference | | | | |
| Yes | **1.36 (1.02, 1.81) *** | 1.06 (0.65, 1.73) | **2.02 (1.31, 3.11) *** | 1.21 (0.63, 2.30) | 1.11 (0.60, 2.07) |
| **Blood level of lead (unit 10 µg/dL)** | **1.60 (1.20, 2.14) *** | 1.74 (0.95 3.20) | **2.38 (1.52, 3.71) †** | 0.45 (0.10, 2.11) | 0.23 (0.03, 5.37) |

Cadmium analysis based on complete dataset with information of blood lead and cadmium measurements with N=42,255. Multiple imputation was performed in univariate and multivariate logistic regression models. Variables with P value < 0.1 in the univariate in total cohort were included into multivariate analysis. *P < 0.05, †P < 0.001, ‡P <0.0001. Abbreviations: odds ratio, OR; confidence interval, CI; body mass index, BMI; Non-Hispanic White, NHW; Non-Hispanic Black, NHB; Mexican American, MA; other race, O.

**Table S8. Odds ratios from multivariable logistic regression models with the outcome of fibrosis defined by FIB-4 >= 2.67 and/or Forns >= 6.9 in no exposures identified group**

| **Cohorts** | **No exposures identified group, OR (95% CI)** | | |
|---|---|---|---|
| **Sample size (N)** | **Total (9,534)** | **NHW (4,372)** | **NHB (2,163)** |
| **FIB-4 >= 2.67 and/or Forns > 6.9 (N)** | **451** | **291** | **87** |
| **Age (unit 10 years)** | **3.21 (2.78-3.70) ‡** | **3.16 (2.62-3.80) ‡** | **4.14 (2.94-5.84) ‡** |
| **Gender** | | | |
| Female | Reference | | |
| Male | **2.29 (1.72, 3.05) ‡** | **2.20 (1.56, 3.09) ‡** | **2.39 (1.32, 4.32) *** |
| **Kidney insufficiency** | | | |
| No | Reference | | |
| Yes | **1.46 (1.08, 1.96) *** | **1.50 (1.06, 2.13) *** | 1.33 (0.72, 2.47) |
| **Diabetes** | | | |
| No | Reference | | |
| Yes | 1.28 (0.91, 1.82) | 1.37 (0.89, 2.10) | 0.80 (0.47, 1.36) |
| **Hypertension** | | | |
| No | Reference | | |
| Yes | 1.29 (0.93, 1.78) | 1.17 (0.80, 1.70) | 1.30 (0.47, 3.61) |
| **BMI** | | | |
| Normal | Reference | | |
| Overweight | 0.91 (0.64, 1.29) | 0.95 (0.64, 1.41) | 0.66 (0.36, 1.21) |
| Obese | 0.82 (0.57, 1.19) | 0.86 (0.55, 1.35) | 0.69 (0.37, 1.30) |
| **Alcohol use** | | | |
| Lifetime abstainers | Reference | | |
| Former drinkers | 1.49 (0.9, 2.45) | 1.55 (0.85, 2.83) | 1.61 (0.69, 3.71) |
| Non-excessive current drinkers | 0.95 (0.6, 1.49) | 0.88 (0.51, 1.49) | 1.44 (0.52, 4.01) |
| **Smoking status** | | | |
| Never | Reference | | |
| Past/current | 1.02 (0.77, 1.34) | 0.96 (0.69, 1.35) | 1.22 (0.70, 2.13) |

Multiple imputation was performed in multivariate logistic regression models. Variables with P value < 0.1 in the univariate in total cohort were included into multivariate analysis. *P <0.05, †P <0.001, ‡P <0.0001. Limited by fibrosis cases, analysis did not provide in Mexican American and other race. Abbreviations: odds ratio, OR; confidence interval, CI; body mass index, BMI; Non-Hispanic White, NHW; Non-Hispanic Black, NHB.
